# Effect of Lockdown Implementation, Environmental & Behavioural factors, Diet and Virus Mutations on COVID-19 Outcomes: A Study on Critical Containment Zones of Indian state of Maharashtra

**DOI:** 10.1101/2020.09.12.20193078

**Authors:** Onkar T. Mohite, Arvind S. Avhad, Prasad P. Sutar, Vaibhav S. Pawar

## Abstract

This work details the study of critical containment zones in Maharashtra within a time (April 9 2020 - July 31 2020) in the context of COVID-19. The effects of lockdown implementation, community isolation, environmental factors, demographic aspects, behavioural factors, diet etc. have been investigated. The effect of the aforementioned factors on the infected cases, cumulative infected cases, recoveries, cumulative recoveries, active cases, deaths and cumulative deaths are analyzed. The integrated effects of the aforementioned factors on COVID-19 outcomes are further amplified due to adequate and inadequate health facilities. The study will be helpful to scientists, researchers, pharmacists and biotechnologists in new vaccine design & to accommodate above factors for the betterment of susceptible & infected people of Maharashtra and similar demographies across the globe. Further, it pinpoints the need for more awareness and control strategies among the people to reduce the havoc, stress, fear, anxiety, pathogenicity and thereby reducing mortality.

## 1. Introduction

The outbreak of novel coronavirus (COVID-19) was initiated with its first reported case on March 9, 2020, in the Indian state of Maharashtra. Since then, the state has been reported with 422,118 confirmed positive cases, 14,994 deceased cases as well as 256,158 recovered cases as of July 31, 2020. One-third of the nation’s total COVID-19 cases have been reported in Maharashtra alone. Considering the same massive rate of spread around the globe, the development process of vaccines by many researchers had begun with the deciphering of the SARS-CoV-2 genome in January 2020. However, there are many uncertainties in the path owing to the possibility of failures in some of the trials, as well as some trials yielding unclear results. With a prolonged period of the time investment involved in the development of the vaccine, we have to wait for a year or so until a safe and effective vaccine is made available to the global community. Due to the same reason, many researchers have begun work in the context of spread patterns, factors affecting the spread, mortality and recovery, as well as the antibody mechanism of this novel coronavirus.

COVID-19 under-reporting problems are addressed by Sezer Kisa et al. [1] through the case study of turkey. A Wilder-Smith et al. [2] highlighted the importance of conventional methods of isolation, quarantine, social distancing and community isolation in the context of COVID-19. L Caly et al. [3] described the isolation and sequencing of COVID-19 patients in Australia. M Day et al. [4] presented a case study of an Italian village where the identification and isolation of asymptomatic cases helped eliminating the virus. K Usher et al. [5] worked modern quarantine strategies for social isolation and studied mental health in the pandemic. The influence of environmental effects on SARS-CoV-2 patients has been evaluated by F Magurango et al. [6]. A Gasparrini et al. [7] studied epidemiologic evidence of environmental and societal determining factors to respond to COVID-19.

Md. Hasan Al Banna et al. [8] explored the impact of the COVID-19 pandemic on mental health among a representative sample of home-quarantined Bangladeshi adults. Giulia Landi et al. [9] examined the mediating and moderating roles of psychological flexibility in the link between trait health anxiety and three mental health outcomes: COVID-19 peritraumatic distress, anxiety and depression. Chiara Baiano et al. [10] presented a valuable approach to support individuals experiencing anxiety related to the COVID-19 outbreak could be represented by mindfulness-based interventions improving the ability to focus attention and awareness on the present moment.

S J de Vlas et al. [11] discussed practical strategies to achieve herd immunities at wider areas with the help of a phased lift of control. HE Randolph et al. [12] explained basic concepts of herd immunity in the context of COVID-19. K Syal et al. [13] worked on herd immunity and convalescent plasma therapy in the context of COVID-19. VB Bulchandani et al. [14] presented insights on herd immunity to epidemics like COVID-19. The work highlights that not all individuals in the community are required to be immune. KO Kwok et al. [15] worked on the estimation of the level of herd immunity required to halt COVID-19 in profoundly affected countries. E slot et al. [16] provided vital information on the extent of the outbreak in countries and herd immunity is not a realistic option for an exit strategy. Eleni P. et al. [17] stated that long term herd immunity might not be sufficient to control COVID-19 and there is an urgent need for an effective vaccine.

Dimple Rawat et al. [18] presented the importance of non-pharmacological approaches like a balanced diet, stress management, adequate sleep and physical activity to build up a robust immune system, as it reduces the complications in individuals who are already at nutritional risk and might get exposed to the infection. Rahul Kalippurayil Moozhipurath et al. [19] empirically outlined the independent protective roles of lockdown and UVB exposure as measured by the ultraviolet index (UVI), whilst also examining whether the severity of lockdown is associated with a reduction in the protective role. Virna Margarita Martín Giménez et al. [20] discussed and proposed whether or not vitamin D and renin-angiotensin-aldosterone system ethnical disparities influence susceptibility to infection and death by COVID-19 in black people & suggested possible mechanisms for this susceptibility. Ranil Jayawardena et al. [21] evaluated pieces of evidence from clinical trials of earlier viral strains in the context of nutrition-based interventions of viral diseases. Fayth L Miles et al. [22] studied the difference in the plasma, urine and biomarkers of dietary intake between vegetarian and non-vegetarian diet groups. Gina S-S. et al. [23] compared the dietary intakes of vegetarian and non-vegetarian individuals concluding that vegetarians have a more favourable intake profile overall. Josefine Nebl et al. [24] compared vegetarian and non-vegetarian dietary patterns for the evaluation of exercise-induced oxidative stress, nitric oxide and plasma amino acid profile in recreational runners. Moshe R. et al. [25] reviewed the antiviral potential of Cepharanthine; a Japanese approved drug in the context of COVID-19. The impact of BCG on prevention of spread and reducing the severity of COVID-19 is studied by AR Sharma et al. [26].

Y H Cheng et al. [27] assessed exacerbated inflammatory effects posed by secondary pneumococcal pneumonia, given prior influenza infection. M Cantone et al. [28] presented a multiplicity of mathematical modelling in the context of bacterial lung infections. Nicholas P Joseph et al. [29] carried out work on the racial/ethnic disparities in disease severity of patients admitted with confirmed COVID-19. Jude Fernando et al. [30] worked on the vulnerability nexus of viruses, capitalism and racism in the context of COVID-19. Ahmed Nabil et al. [31] presented a review on coronavirus in the context of epidemiological, diagnostic and therapeutic approaches. Wenrui Hao et al. [32] developed a mathematical model based on the interactions among cells and proteins that are involved in the progression of the disease. The model simulations are shown to be in agreement with available lung tissue data of human patients. Babak Eshratithe et al. [33] studied factors affecting the survival rate of COVID-19 patients in Iran using a retrospective cohort. Esteban Correa Agudelo et al. [34] characterized vulnerable populations located in areas at higher risk of COVID-19 related mortality and low critical healthcare capacity in the US: using Bayesian multilevel analysis and small area disease risk mapping. Christopher Leffler et al. [35] determine sources of variation between countries in per-capita mortality from COVID-19 (caused by the SARS-CoV-2 virus).

This work is aimed at conducting a thorough study of critical containment zones of Maharashtra within a stipulated time (April 9 2020 - July 31 2020). In the context of the same, the effect of demographic aspects, psychological behaviour, environmental factors, diet, community isolation, lockdown implementation, etc. have been investigated.

The effect of the aforementioned factors on the infected cases, cumulative infected cases, recoveries, cumulative recoveries, active net cases (active cases), and deaths are detailed.

The following are the objectives of the paper with particular focus on COVID-19 outcomes:-

> (a)To carry out the critical analysis of infected cases, cumulative infected cases, recoveries, cumulative recoveries, active net cases (active cases), deaths, and identification of appropriate mathematical functions to mimic the behaviour; (b)To investigate the effect of restricted (controlled) mutation and unrestricted (uncontrolled) mutation in containment zones under study; (c)To investigate the effects of psychological factors such as anxiety, stress, fear, diet, and comorbidities; (d) Addressing the effect of adequate as well as inadequate medications of corresponding health facilities; (e) To investigate the effects of environmental factors; (f) To investigate the integrated effects of the factors mentioned above, in particular on mortality; (g)Helping government sources, doctors, healthcare professionals, scientists, researchers, to frame control strategies and in vaccine design; and (h) To investigate the wavy nature of COVID-19 outcomes.

## 2. Methods

In this study, effects of lockdown implementations, environmental factors, community isolations, psychological behaviour, herd immunity mechanisms, dietary patterns, mortality are analyzed in the context of COVID-19 across containment zones. For this purpose, ten containment zones across the state of Maharashtra, as shown in Fig.1 declared by state authorities are taken into consideration. Data is collected for the individual containment zones from April 9 2020 to July 31 2020, through district information offices of respective areas under consideration, as shown in Table 1. Data post-July 31 2020, is not considered due to the high under-reporting and over-reporting of cases (Data of deceased cases for Mumbai GN and Mumbai ME zones are not taken as segregated data for individual zones in Mumbai is not available). As four containment zones in Mumbai Metropolitan Region (MMR) are considered, zones in Pune district are purposely not included and instead other similar zones which are at higher distance from Mumbai are considered.

**Fig. 1.**
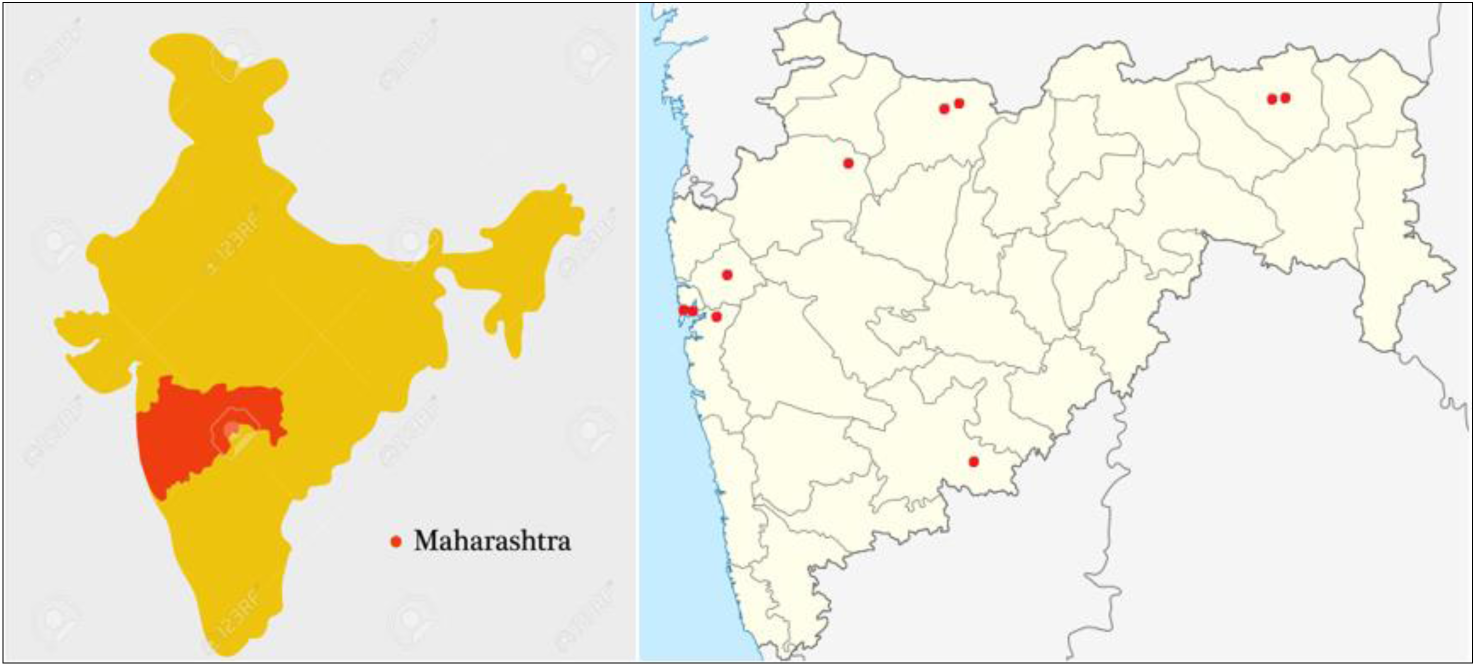
Containment zones under consideration.

**Table.1.** Sample of data collection (Source: DIO Maharashtra) [36] [37].

| DATE | DAILY POSITIVE | DAILY DEATHS | DAILY RECOVERY | ACTIVE CASES |
| --- | --- | --- | --- | --- |
| 09/04/2020 | 5 | 1 | 0 | 4 |
| 15/05/2020 | 14 | 0 | 0 | 497 |
| 30/06/2020 | 0 | 1 | 8 | 141 |
| 31/07/2020 | 9 | 0 | 9 | 93 |

Data obtained through government sources is first analyzed graphically to obtain the best possible fits (R-square) against the lockdown implementations and environmental factors. Further, other factors viz. psychology, viral genetics and dietary patterns are studied in the context of the social demography. The study of mortality is essential to strive for better public health in this terrible viral outbreak of novel coronavirus. This is evaluated via a cumulative deaths over cumulative period (in the batch of 10 days cumulatively i.e. 10 days, 20 days, 30 days and so on from beginning period of study till the period of interest). In the end, all the above-mentioned factors are analyzed in view of cumulative deaths. The flowchart for the present work can be summarized, as shown in Fig.2.

**Fig. 2.**
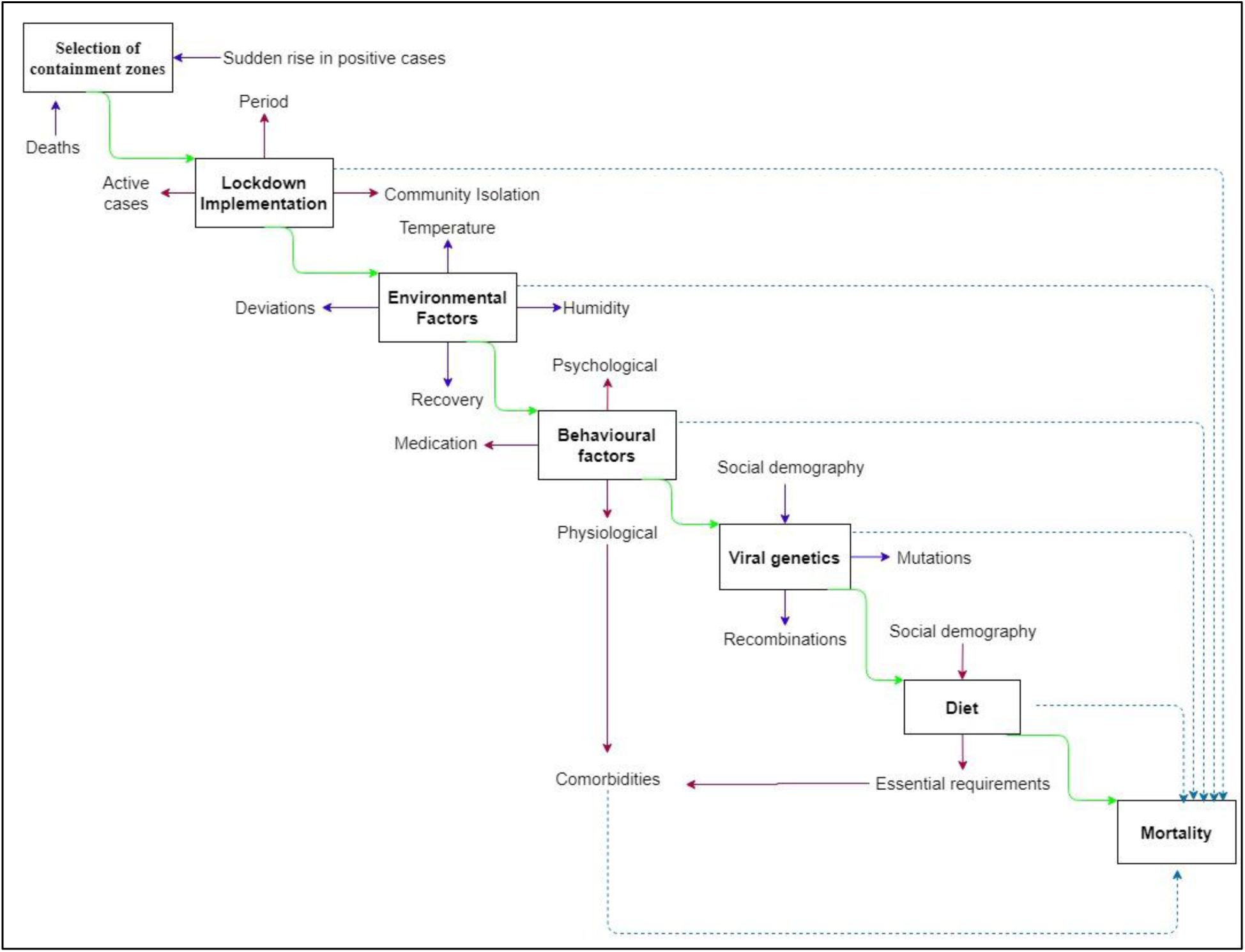
Flowchart for current work.

Statistical analysis of lockdown implementation and environmental factors is discussed in the first two sections. It is then followed by the effects of behavioural factors in the context of psychology, physiology and their relationship with comorbidities. The next section describes social demography and viral genetics (controlled, uncontrolled mutations and recombination). Dietary pattern diverseness in the light of demographic variations and its effects on comorbidities are discussed in the further segment. In the end, cumulative deaths (cum. Deaths hereafter) for each containment zone considered are analyzed and the effects of all aforementioned factors on same are evaluated.

## 3. Results and discussion

Quantitative observational data collected for ten containment zones are evaluated graphically to study the spread pattern of the virus, the ability of a socially isolated community to adopt the changed conditions during and after COVID-19. Also, the effects of lockdown implementation, environmental factors, the behavioural factors of communities in these high-risk containment zones, are evaluated.

### Lockdown Implementation

Malegaon, which is the largest containment zone in Maharashtra, has a population of nearly 12.5 lakhs and 69 km2 total area. 1307 total number of patients have been tested positive for COVID-19, and 1129 patients entirely recovered in Malegaon as of July 31, 2020. The total number of deceased cases reported in Malegaon is 85. The state government had imposed strict lockdown with new laws and enforcement since beginning of May 2020, considering the surge in positive cases, as shown in Fig.3. The highest number of positive COVID-19 cases, i.e., the peak of positive cases, was observed on May 5, with 101 suspected cases tested positive in a single day.

**Fig.3.**
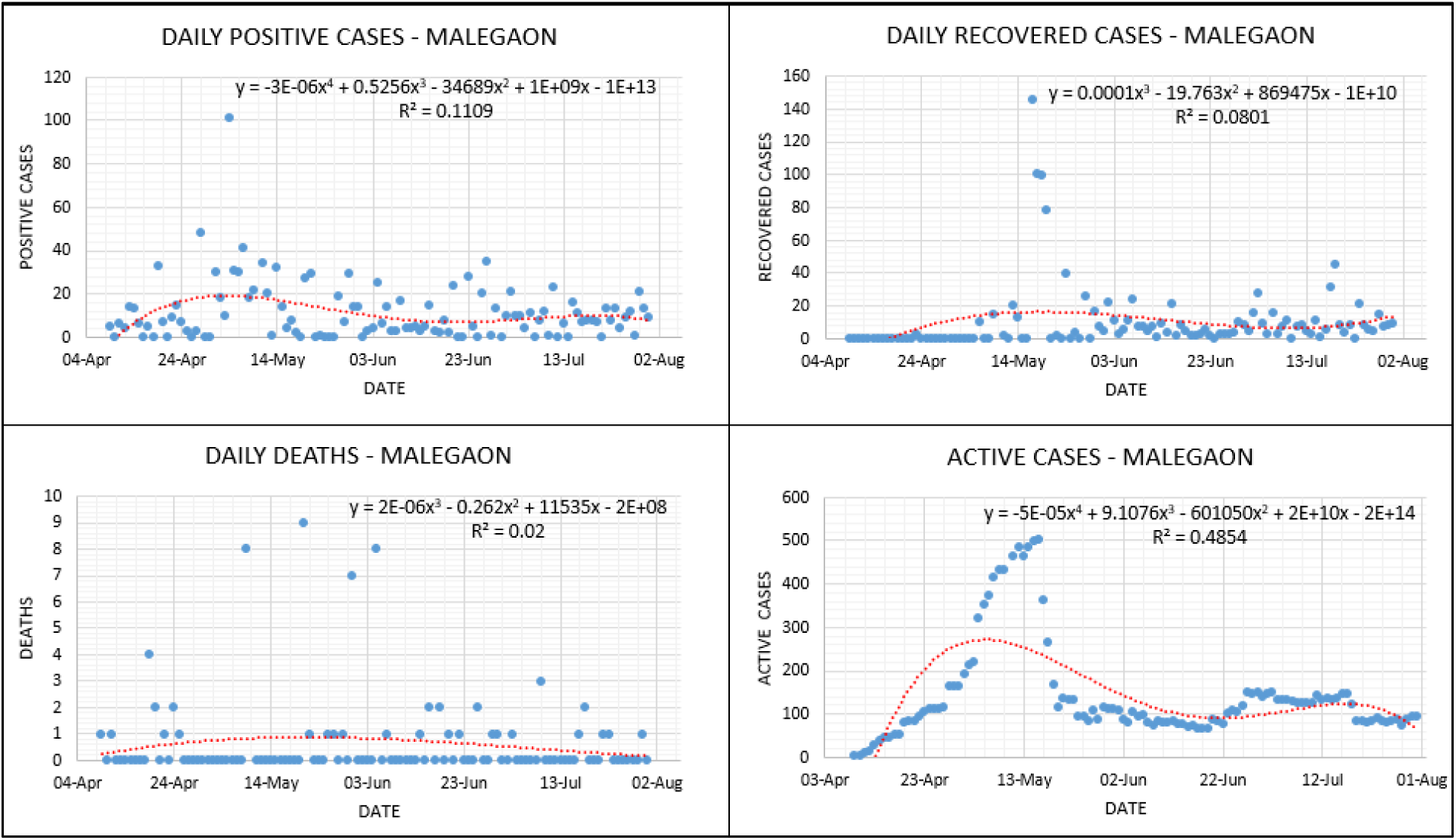
Lockdown Analysis – Malegaon MNC.

The gradual infection spread before the peak is observed suggests that positive cases were gradually increasing until the number of days required for doubling positive cases dropped drastically. The number of cases recovered daily started significantly increasing, as shown in Fig.3, after the peak of infection is observed. After 12 days, the highest number of daily recovered cases were observed on May 17. During this peak to the peak period, it is observed that the infection spread is decreasing gradually, as shown in Fig.3. By the end of May 2020, strict restrictions on movements of people, goods, and social life were relaxed gradually up to a certain extent considering the number of active cases, as shown in Fig.3. Even with the ease in the restrictions, new positive cases and active cases continued to reduce till mid of June 2020. But owing to panic created by some localized peaks of new positive cases reported, restrictions in some hotspot areas were again made strict, which resulted in some amount of anxiety. Due to this abrupt change in the regulations, positive cases reported as well as active cases, can be seen increasing post-July 15, as shown in Fig.3. At the same time, daily recovered cases are also reducing significantly.

Sataranjipura, one of the worst affected COVID-19 hotspots in Nagpur, had reported in a total of 704 positive cases as of May 15. Considering the rapid increase in the number of positive cases, the government imposed strict restrictions in this area and declared Sataranjipura as a containment zone. From the data collected and analyzed, as shown above, Fig.4. For the containment zone period of Sataranjipura, it is visible that the number of positive cases reported each day had been restricted to single digits after the containment zone declaration. The same is the case for active cases as numbers began to drop with periodic batch recoveries observed significantly. So far, 8 deceased cases have been reported from May 15 to July 31. Mominpura, in similar conditions to Sataranjipura, another hotspot in Nagpur city, which had reported 276 positive cases till May 15, became a containment zone with heavy restrictions on movements. It can be seen that the number of positive cases started decreasing with a significantly higher number of recovered cases (daily reported), as shown in Fig.5. Eventually, active cases were reduced exponentially. Following the restrictions of a containment zone, Mominpura had succeeded in reaching zero active cases mark on June 30, with limiting deceased cases to 5. Comparison between Fig.4 and Fig.5 reveal that, even for a same city, active cases follow different curves. It is to be noted that, wavy nature continued almost throughout as is evident from Fig.3, Fig.4 and Fig.5.

**Fig.4.**
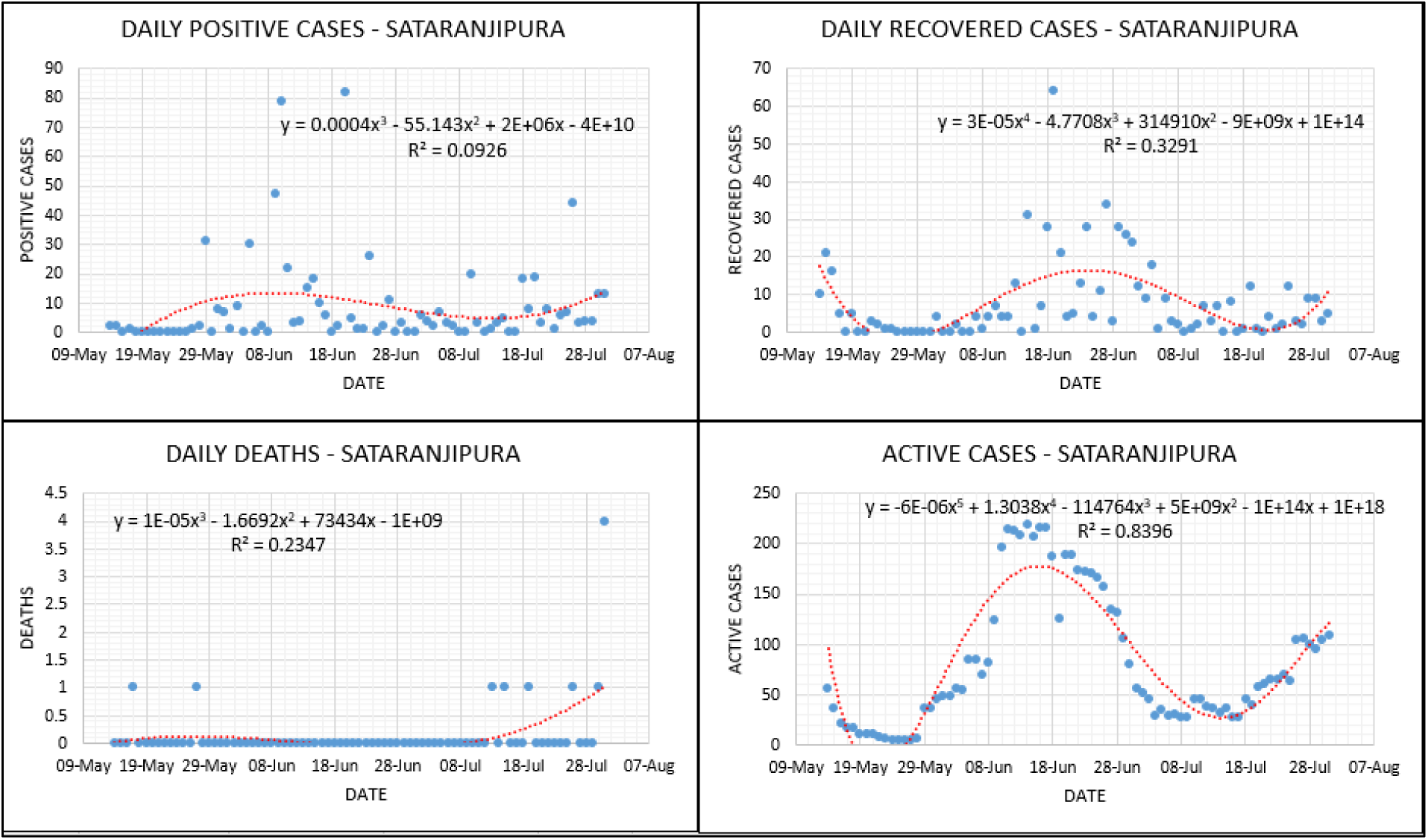
Lockdown analysis – Sataranjipura (Nagpur MNC)

**Fig.5.**
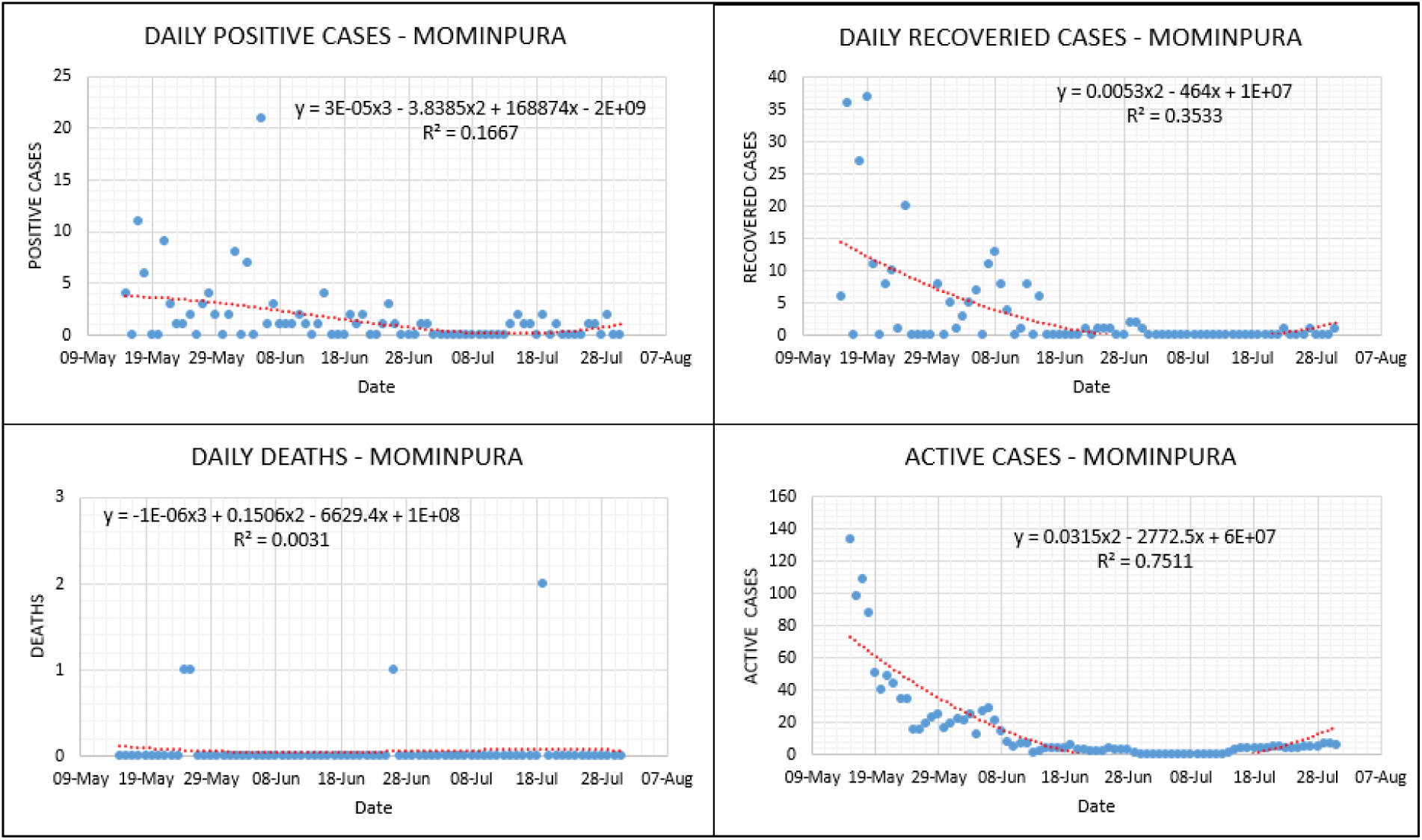
Lockdown analysis – Mominpura (Nagpur MNC)

Mumbai GN ward consisting of Dharavi, Mahim and Dadar and Mumbai ME ward consisting of Mankhurd, Govandi, Deonar, Shivajinagar and Anushakti Nagar are the two of the most high-risk containment zones in Mumbai, which is the worst affected city in India. Graphically these zones are represented in following Fig.6 and Fig.7. Data collected for these two zones is of the post lockdown relaxation period in June 2020. Even though the government lifted strict regulations in other parts of the city, these two most sensitive areas were in continuous monitoring by the concerned authorities. Ease in the restrictions for these high-risk zones was allowed up to a certain extent, only within the boundaries. High fluctuations are observed in the number of daily reported positive cases, but some of the peaks in daily recovered cases are observed in both containment zones. The overall effect can be summarized as though there is no significant reduction in daily positive cases, but due to some mass recoveries, active cases in both zones are decreasing considerably. It can be observed that fluctuations are typically wavy if navigated through scatter. Further, it is clear that in large and densely populated areas like MMR, owing to highly diversified population, outcomes will be fairly complex and therefore outbreak will not vanish soon but will be faded away gradually by wavy path. Very low values of R-square for Fig.3 through Fig.7 for some subplots are due to highly variable nature of pandemic, underreporting, over reporting, tracing problems etc. Further, for the outbreak such as COVID-19, only lockdown and its implementation are not solely responsible for outcomes but other factors do.

**Fig.6.**
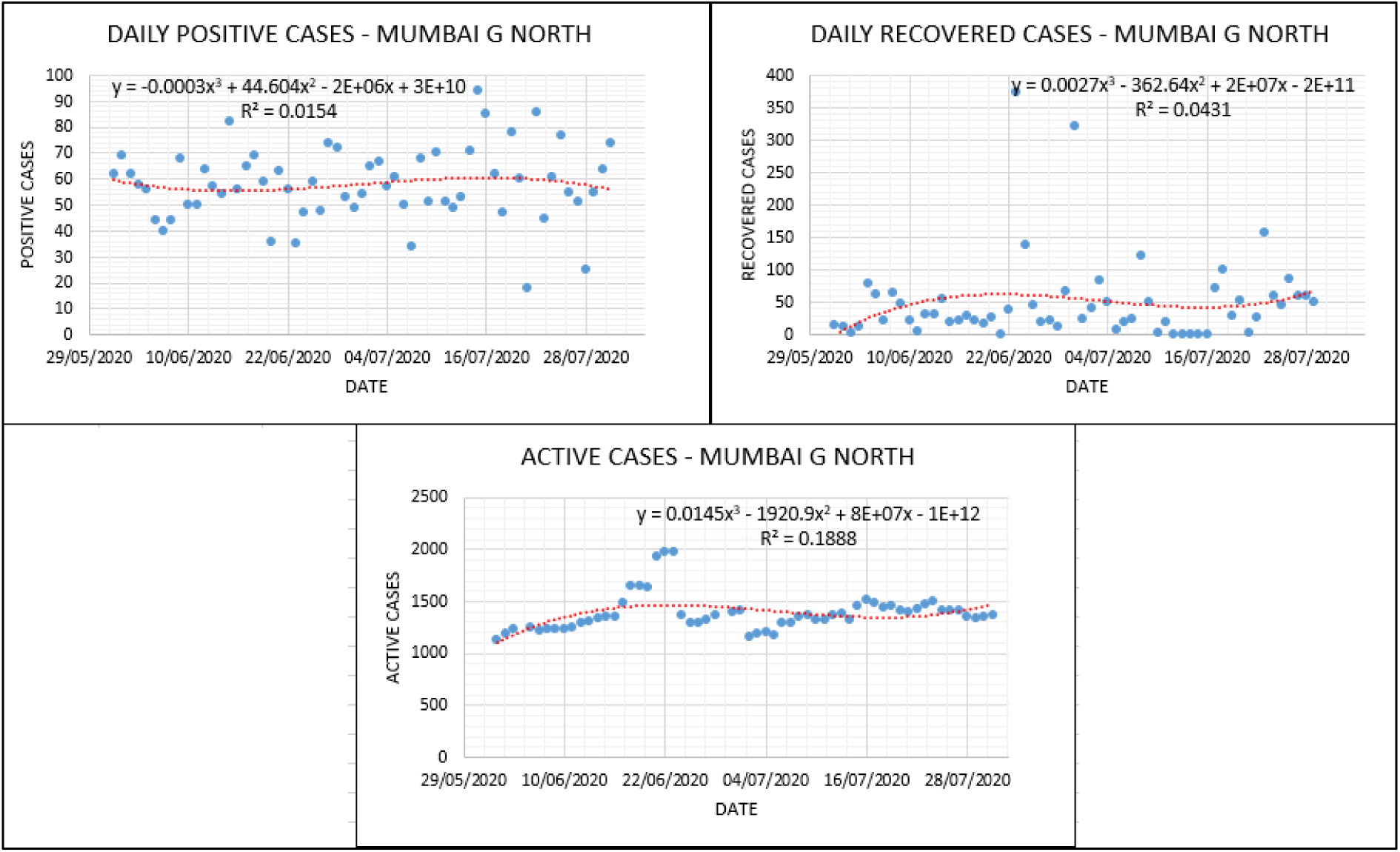
Lockdown Analysis – Mumbai GN ward.

**Fig.7.**
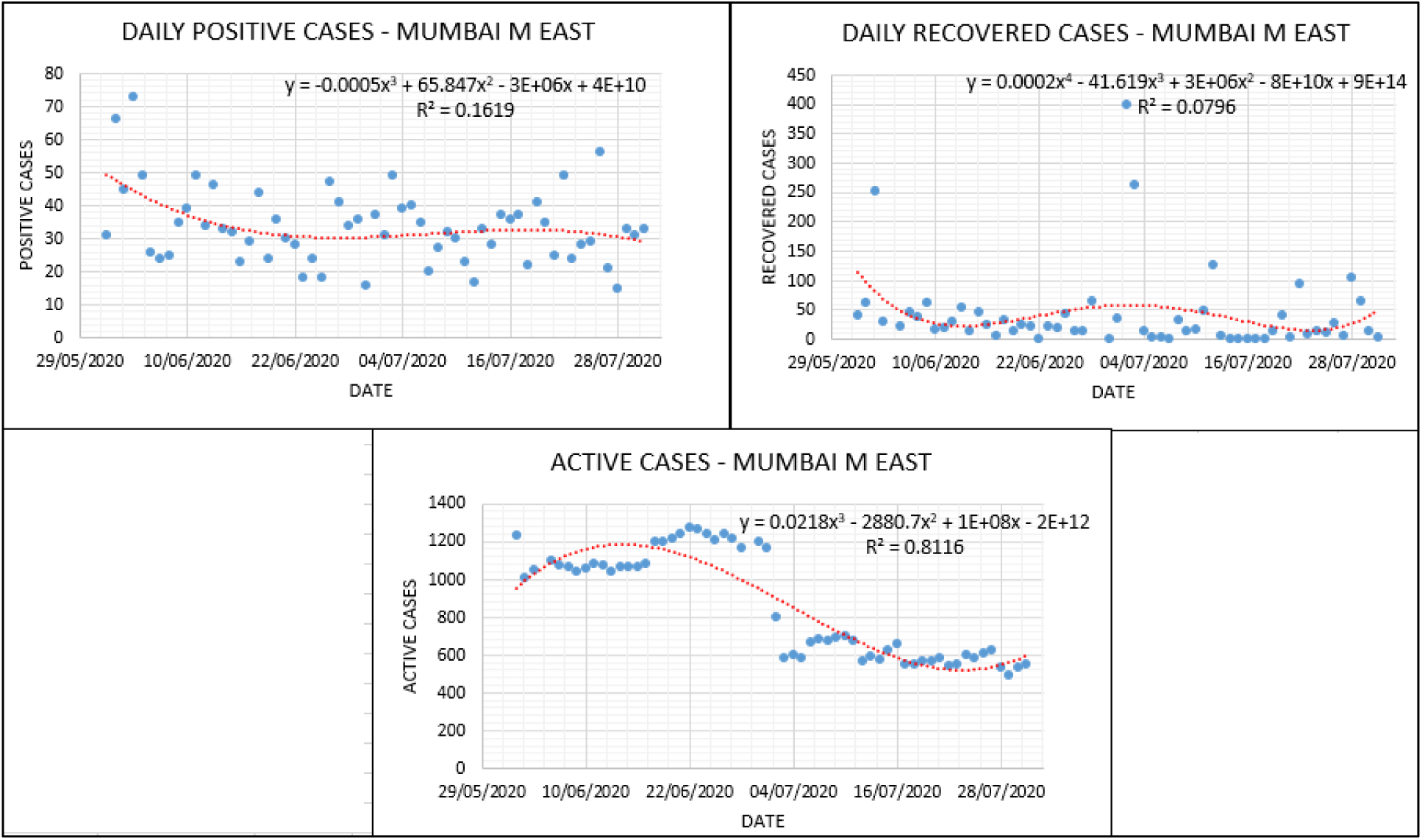
Lockdown analysis – Mumbai ME ward.

Kamothe residency in Panvel Municipal Corporation can be considered as the most well planned and advanced urban developed containment zone in MMR. Similar to the other profoundly affected areas, Kamothe was also declared as a containment zone in the first week of May 2020. Since then, as shown in Fig.8, daily positive cases and active cases can be seen stabilizing slightly throughout containment zone time duration. At the same time, significant batch recoveries are observed in June 2020. Considering this, the strict restrictions were lifted off for a short duration. But as soon as the restrictions on the movements were again imposed in the second half of June 2020, by local authorities, there is an increase in the positive cases (daily reported) and active cases. So far, 38 deceased cases have been reported till the end of July 2020. An increase in deceased cases can be seen post restrictions reinstating period.

**Fig. 8.**
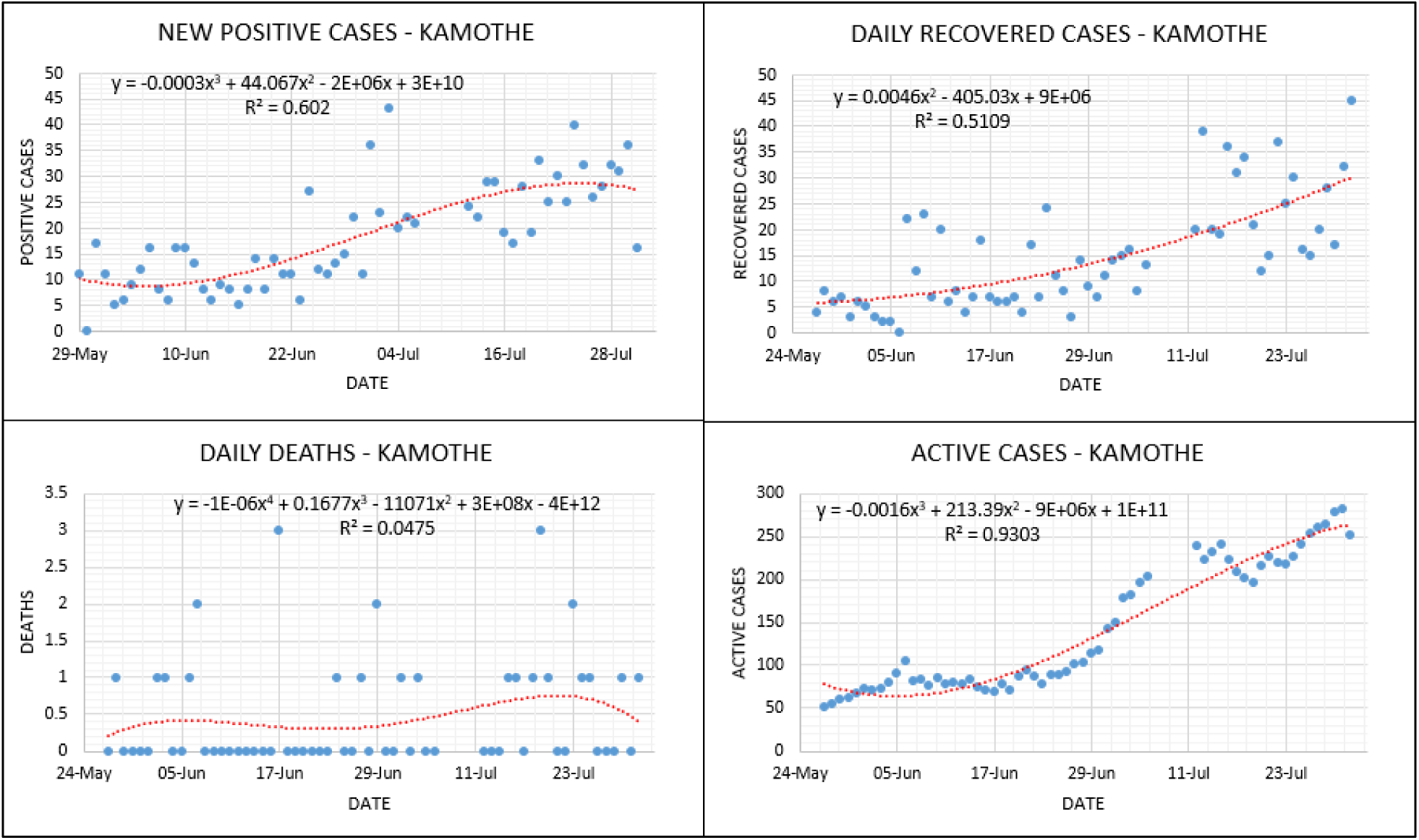
Lockdown analysis – Kamothe.

KDMC is one of the worst affected containment zones (among the zones considered under present work) in terms of the number of positive cases. By the end of July 2020, the total number of positive cases reached 19967. With 357 deceased cases, a total of 13840 patients successfully recovered from COVID-19. As shown in Fig.9 throughout the lockdown period, daily positive cases, daily recovered cases, daily deceased cases reported as well as active cases were increasing continuously until lockdown restrictions were lifted on a much greater extent post-July 15th. Further, one more salient observation is that as positive cases started to increase, nearly in the same period, recovered cases increased. This clearly highlight’s that active cases will increase gradually with not that much steep slopes immediately as is evident From Fig.9

**Fig. 9.**
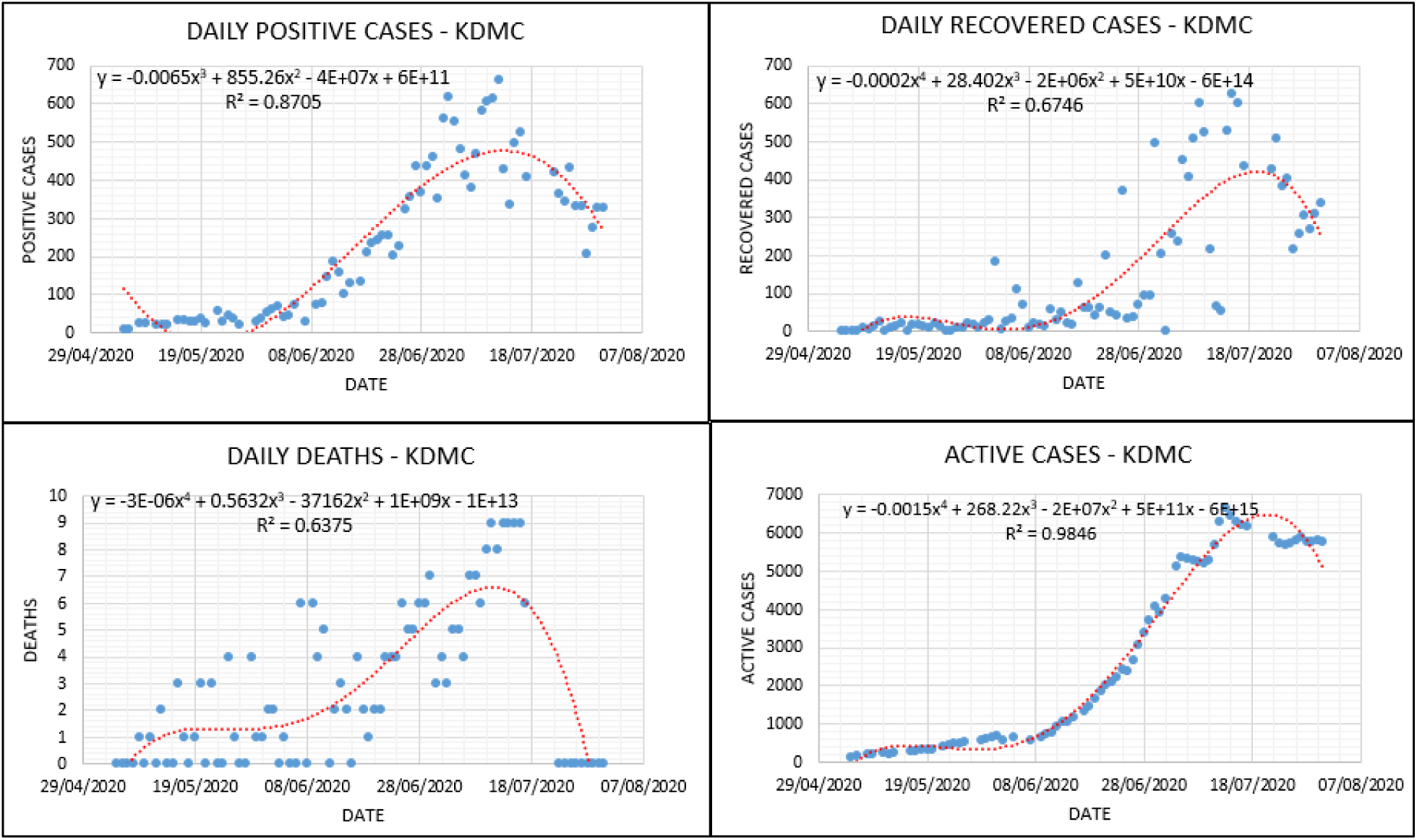
Lockdown analysis – KDMC.

Jalgaon MNC and Bhusaval are the two profoundly affected zones in the northern parts of Maharashtra. In cases of Jalgaon MNC, all four parameters viz. positive cases, recovered cases, deaths, as well as active cases are increasing throughout the lockdown period. However, the increase in daily positive cases is much higher than the number of recovered cases. Hence the curve for active cases can be seen continuously climbing, as shown in Fig.10, with moderate slopes. On the other hand, Bhusaval zones possessed the mixed nature of the curve in the lockdown period. For June 2020, the daily positive cases can be seen decreasing, as shown in Fig.11, which also resulted in stabilization of the net active cases curve in the same period up to a certain extent. Deceased cases are found to be decreasing but by wavy manner. Further, slope patterns for Fig.10 and Fig.11 are different (for instance active cases) as other factors may have significant contributions over COVID-19 outcomes.

**Fig. 10.**
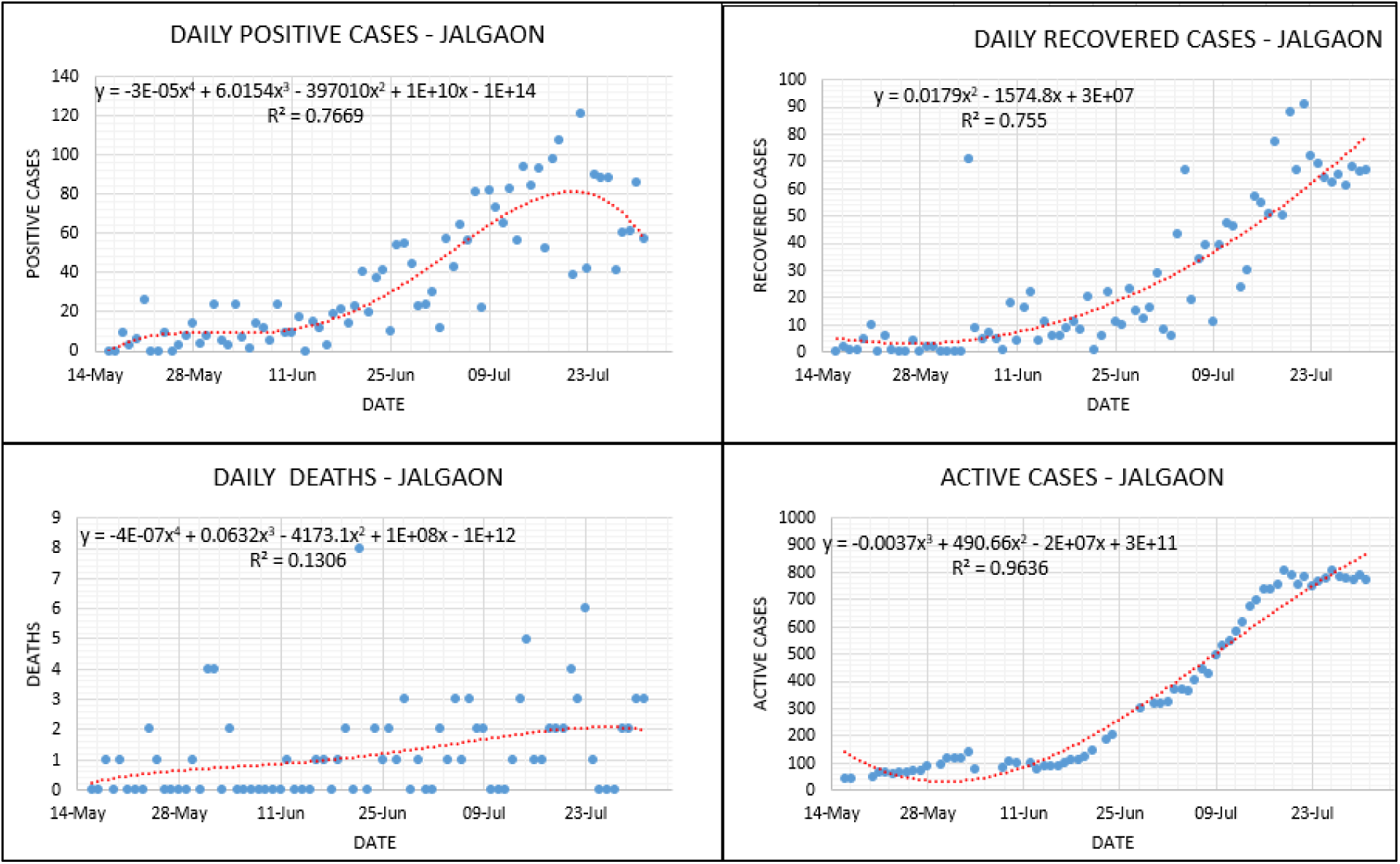
Lockdown analysis – Jalgaon MNC.

**Fig. 11.**
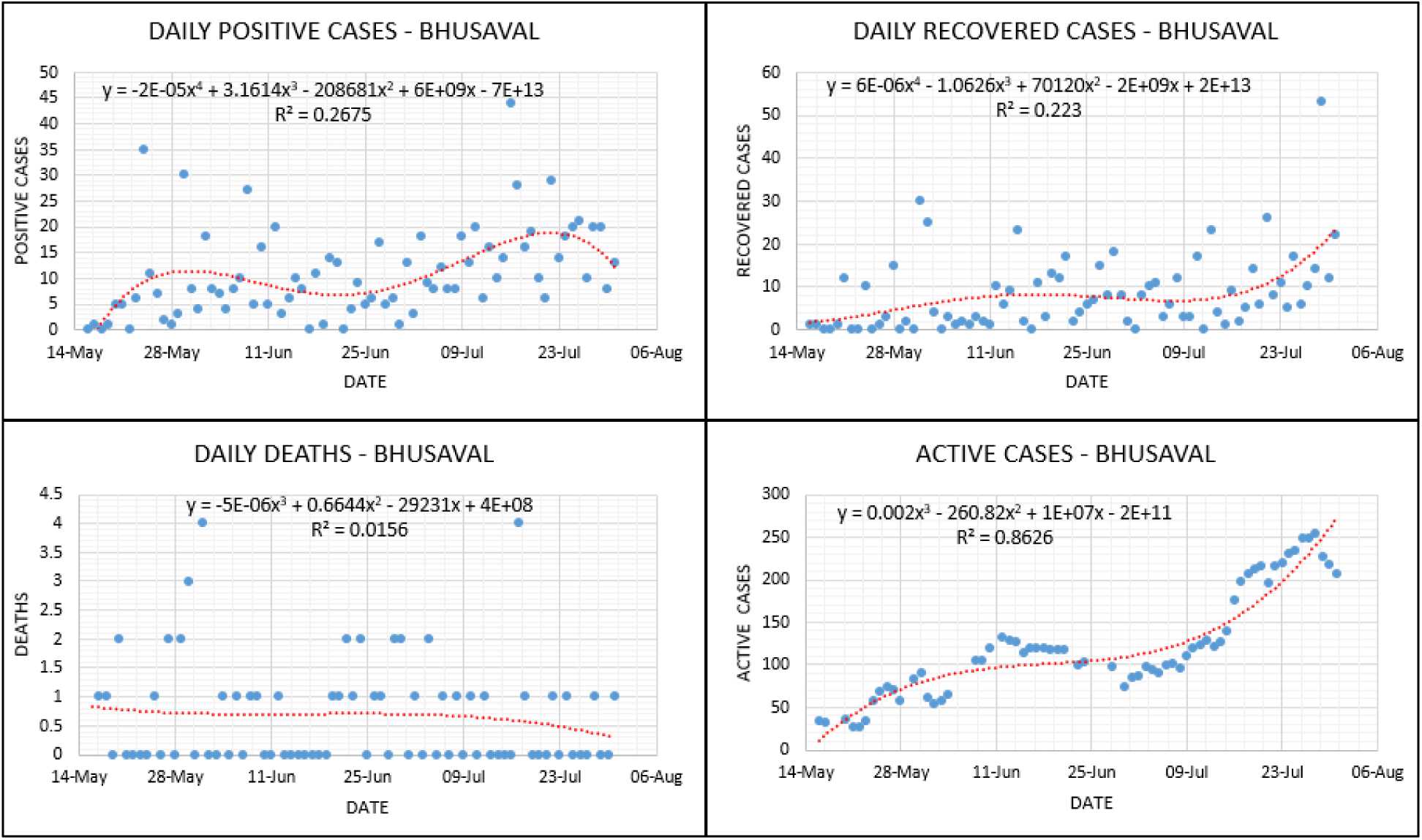
Lockdown analysis – Bhusaval.

Solapur MNC saw a sudden increase in the active cases since second week of June 2020. The main reason behind this may be the slow recovery as compared to the increase in new positive cases, as shown in Fig.12. Deceased cases were found to be fluctuating with large variance (due to wide scatter), causing a total of 351 deaths till July 31, 2020. Other factors might have dominant effect for Solapur which needs critical attention. This region may face secondary or tertiary peaks for the COVID-19 pandemic.

**Fig. 12.**
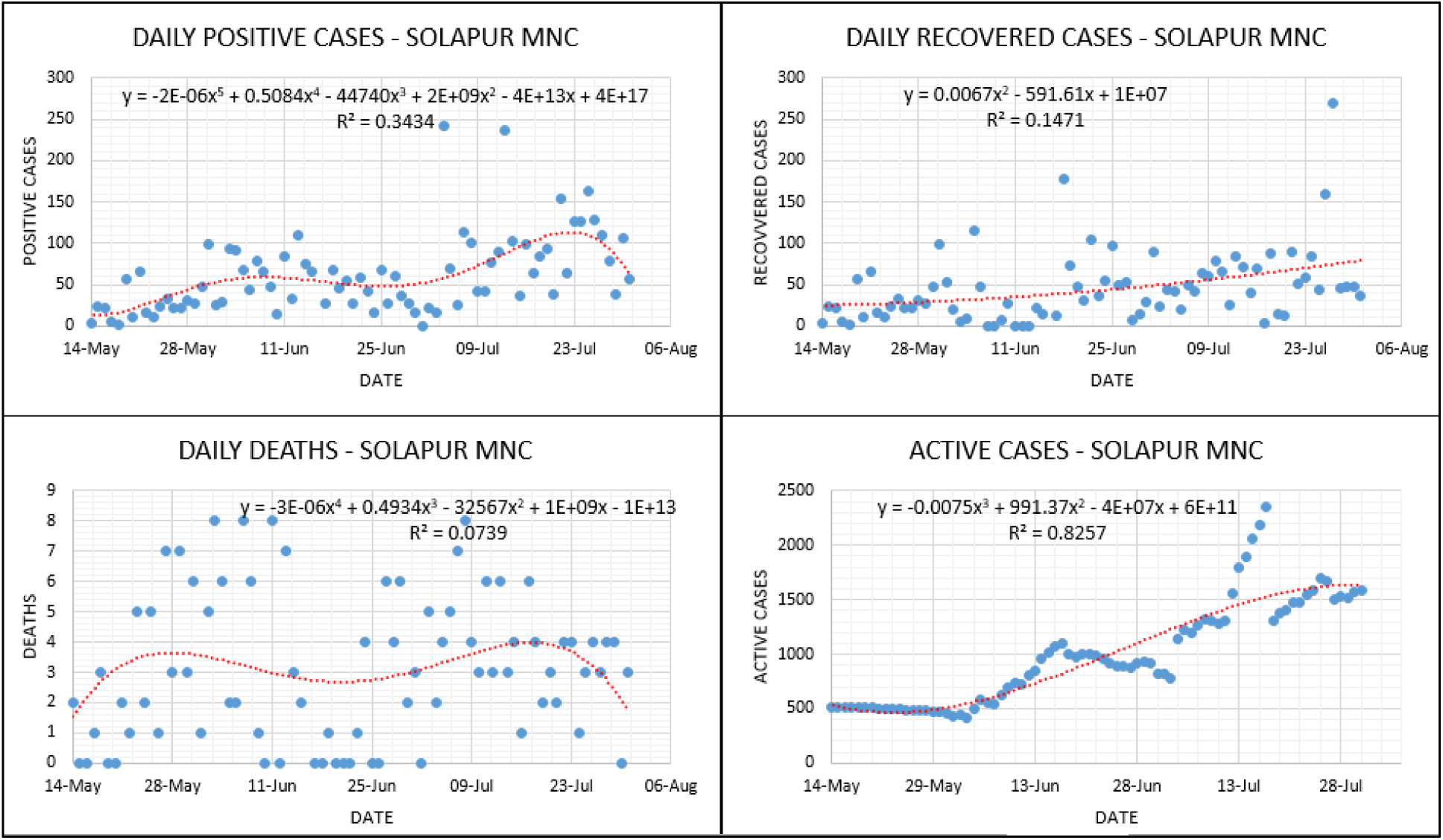
Lockdown analysis - Solapur MNC.

The comparison of all the above-mentioned containment zones shows that the infection spread, recovery as well as deaths in the case of this novel coronavirus, largely depends upon how a group of people in terms of communities are allowed to behave in a pandemic. Malegaon, Sataranjipura, and Mominpura containment zones can be considered as socially isolated communities during the time duration when strict restrictions were imposed. As a result of only exceptional in and out movements in these zones, the infection spread can be seen decreasing with a significant rise in recovered cases reported per day. On the other hand, the remaining containment zones under consideration in MMR show different results. It is crucial to consider the importance of physical boundary separation of a particular area or zone in the context of containment, as earlier three zones possess excellent isolation characteristics. At the same time, regions in MMR have difficulties in maintaining norms of socially isolated communities. As a result, it is seen that the extent up to which the outbreak is controlled in Mumbai GN (excluding Dharavi), Govandi / Deonar / Mankhurd, Kamothe as well as KDMC containment zones are much lower than the earlier three zones. Also, too much attention has been given to the Dharavi area by local governing authorities. Efforts are well appreciated by World Health Organization (WHO). Increased positive cases in other parts GN ward were reported. There is a need to take a note of disease tolerance of the population for other parts of GN ward, unattended population of other similar wards, unattended sections of containment zones & areas nearby containment zones.

From the statistical point of view, R-square values for daily reported data are not significant. Lack of management, under-reporting, over-reporting, other factors as well as natural phenomena etc. might be the possible reasons behind the high variance in the data recorded daily. However, the same data, when analyzed on a cumulative basis, yielded significant values of R-square. For brevity, three zones are analyzed and represented considering cumulative data, as shown Fig.13 through Fig.15. Cumulative data fits, captured the behaviour reasonably well. This is due to fact variations are adjusted in the cumulative data. Nevertheless, for the pandemic such as COVID-19, history of; infections, recovered cases, deaths and active cases are essential while determining outcomes on particular day.

**Fig.13.**
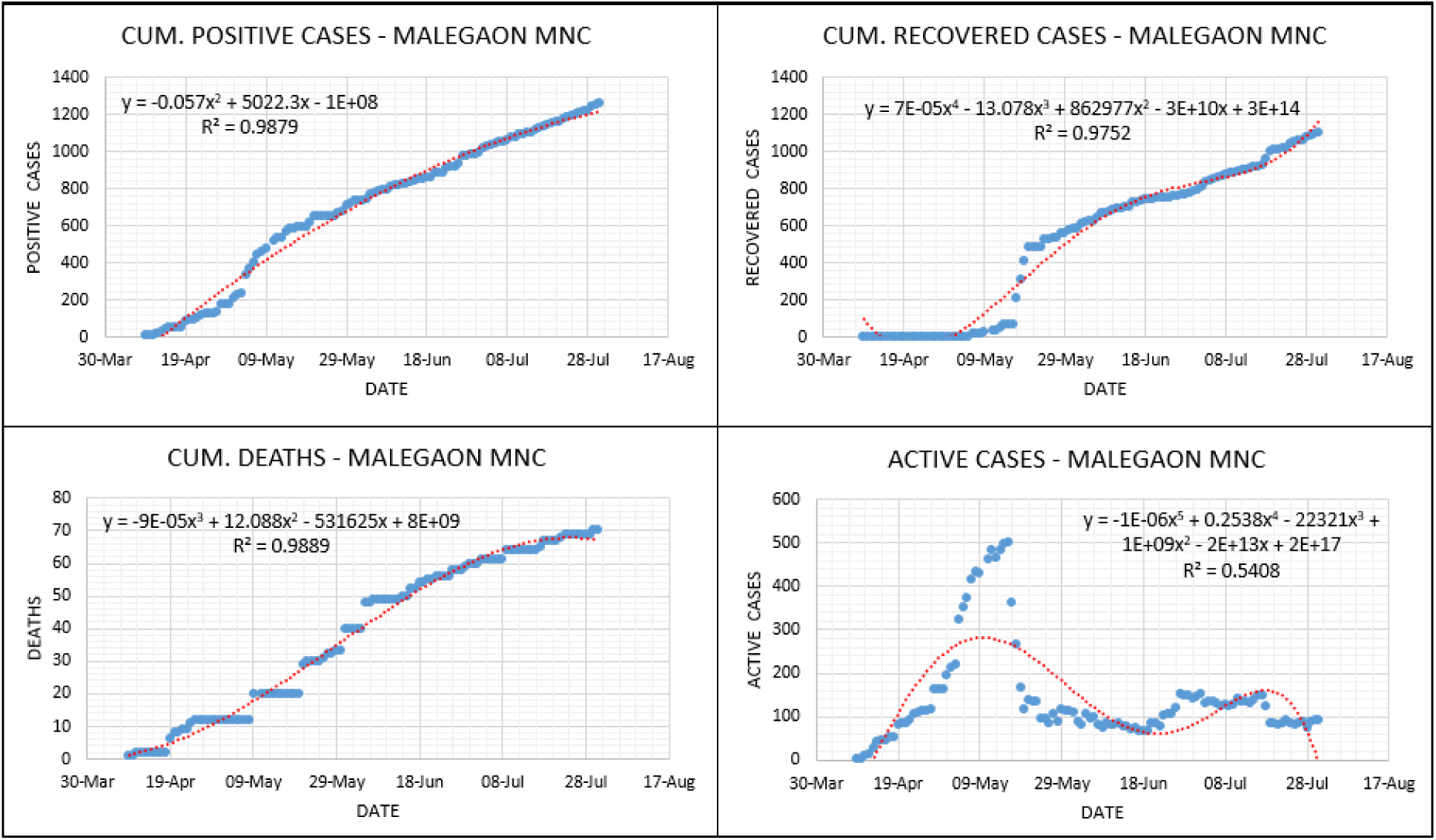
Lockdown analysis cum. data – Malegaon MNC.

**Fig.14.**
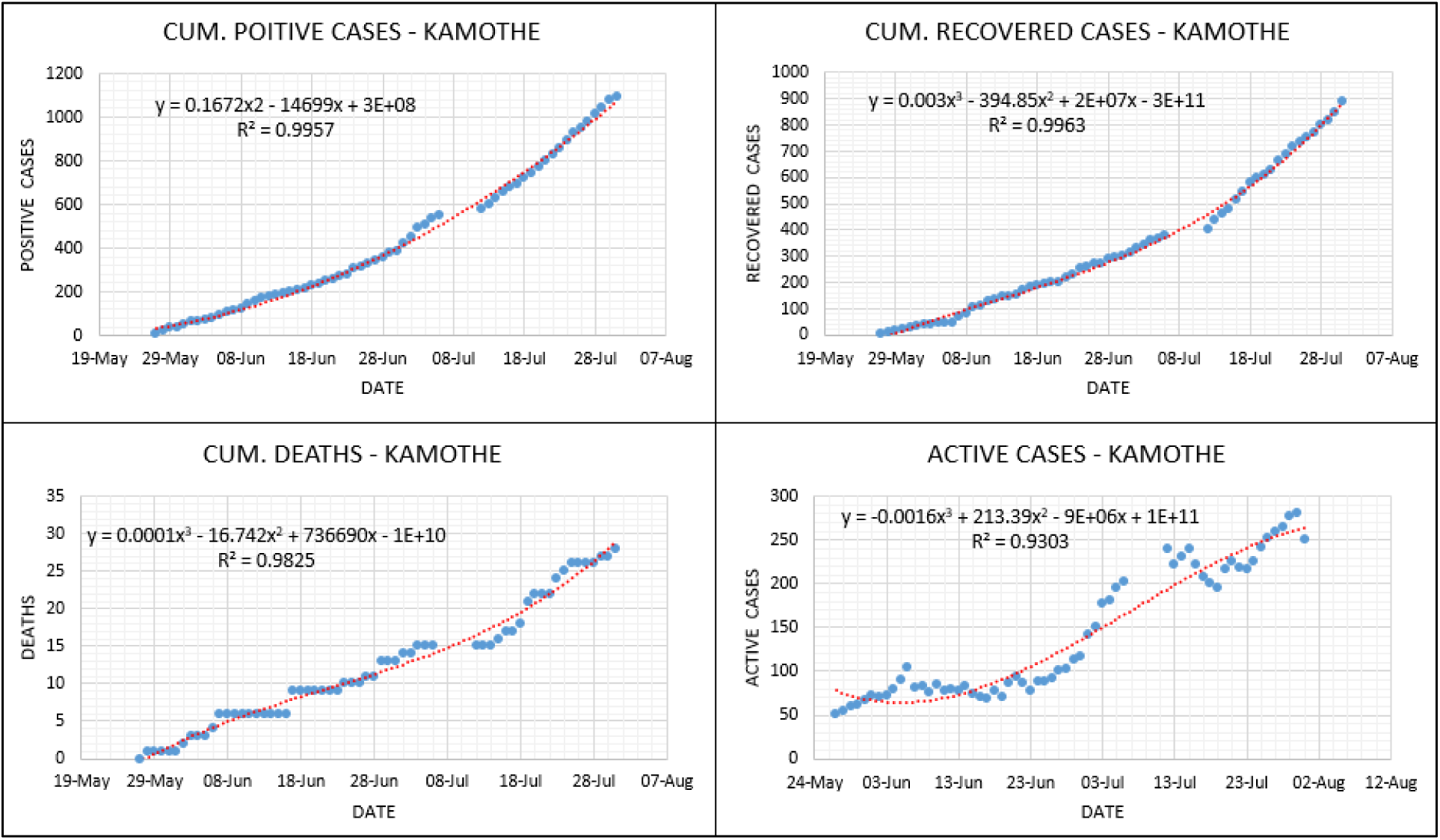
Lockdown analysis cum. data - Kamothe.

**Fig.15.**
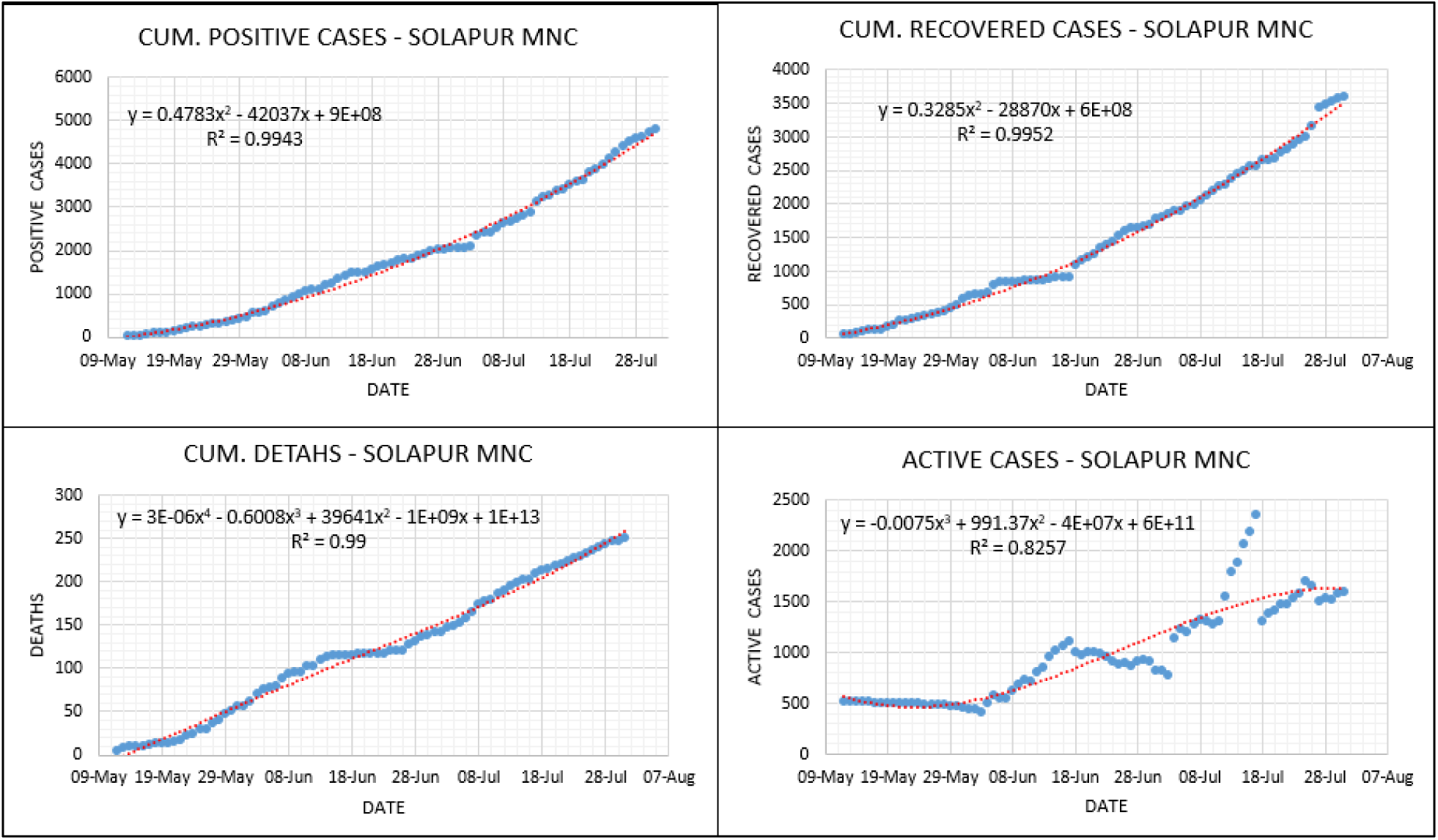
Lockdown analysis cum. data – Solapur MNC.

Too many fluctuations in the policies have resulted in more havoc. Furthermore, this hampered the mental health of the common public severely. Poor medication in most of the rural segments of containment zones worsened things further. On the other hand, almost for all the containment zones waviness in the context of positive cases, recovered cases, deaths and active cases have been observed as evident from Fig.3 through Fig.15. It is to be noted that, in the large population of demographics such as Maharashtra, aforementioned aspects attract attention and need to be reviewed and reformed.

### Environmental Factors

Along with the lockdown imposition and its effects, metrological parameters too play a vital role in the transmission pattern of this novel coronavirus. In the same context, it is essential to evaluate these factors viz. humidity and temperature with the daily recovered cases in all the containment zones under consideration. Correlation of daily recoveries is evaluated separately with mean temperature, temperature deviations, mean relative humidity, and deviations observed in relative humidity, as shown in Fig.16 through Fig.25. R-square values are relatively low for the majority of the containment zones except for Mominpura, Jalgaon MNC, and KDMC, which show statistically significant values of R-square. The reason behind these low values of metrics is high variance in data of the daily recoveries and their over as well as under-reporting. However, when the same data is considered as cumulative and analyzed against environmental factors, better values of R-square are obtained. For brevity, graphs of three zones are shown in Fig.26 through Fig.28. These things can be explained on similar lines as discussed in the previous section.

**Fig.16.**
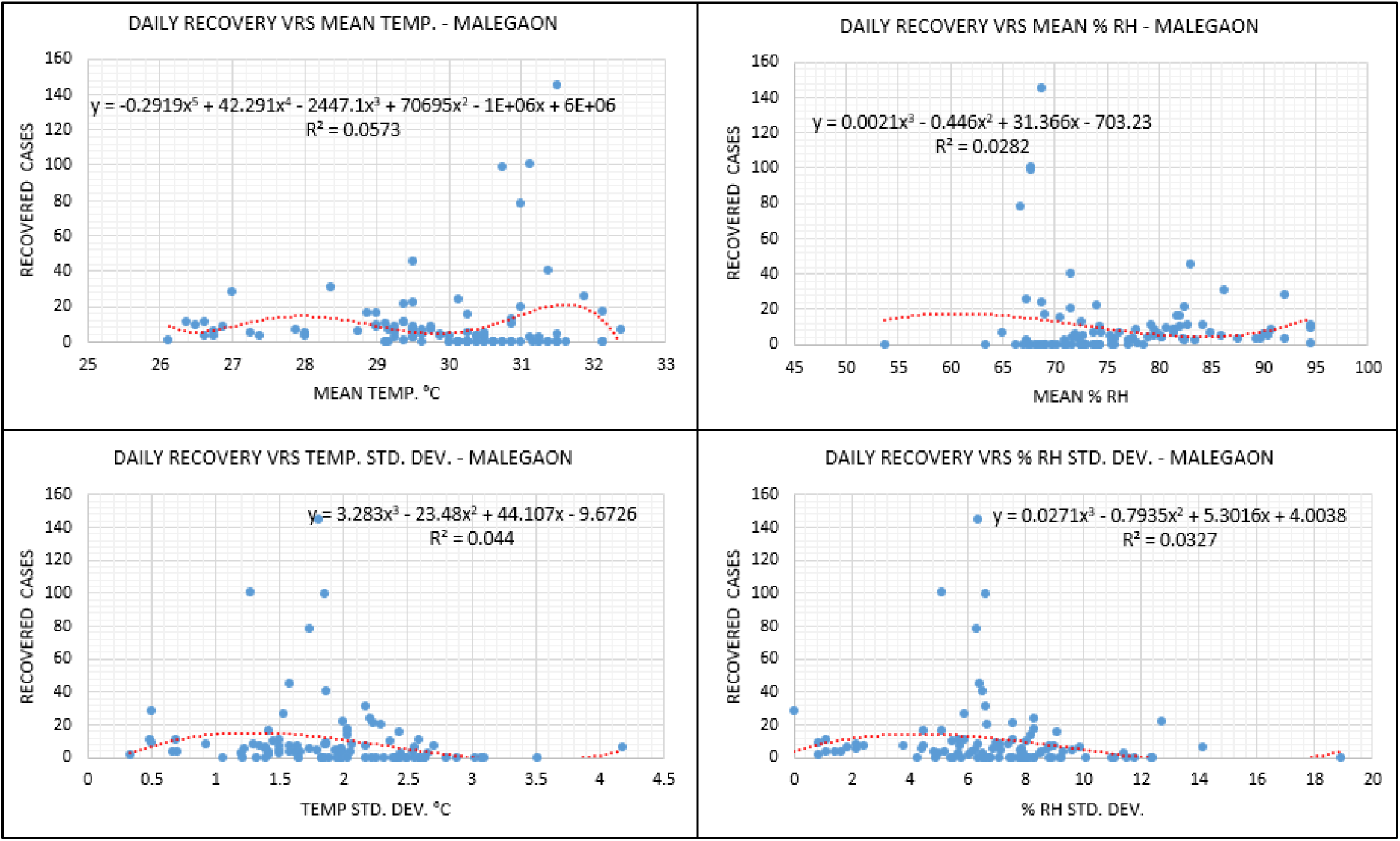
Environmental analysis – Malegaon MNC.

**Fig.17.**
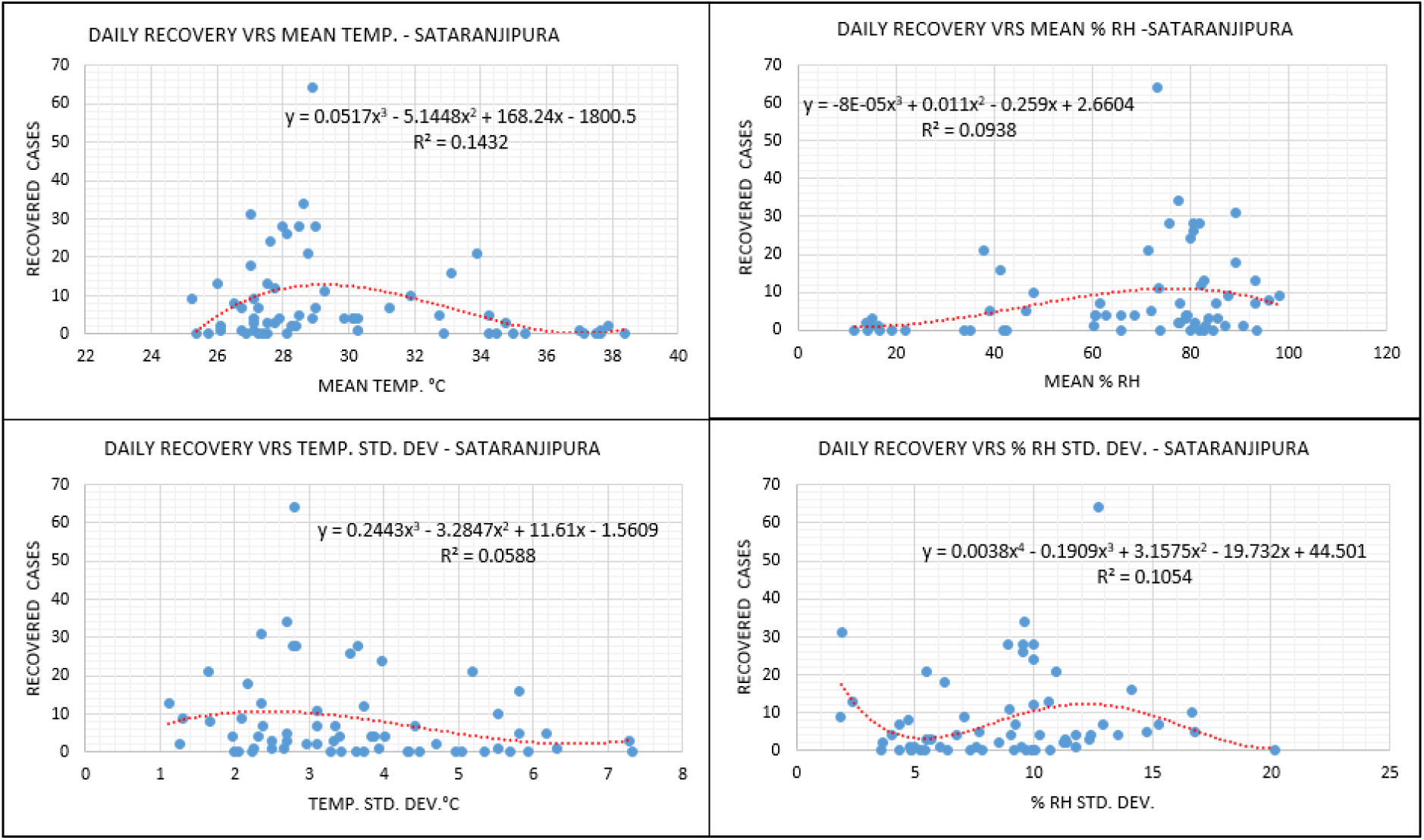
Environmental analysis – Sataranjipura (Nagpur MNC)

**Fig.18.**
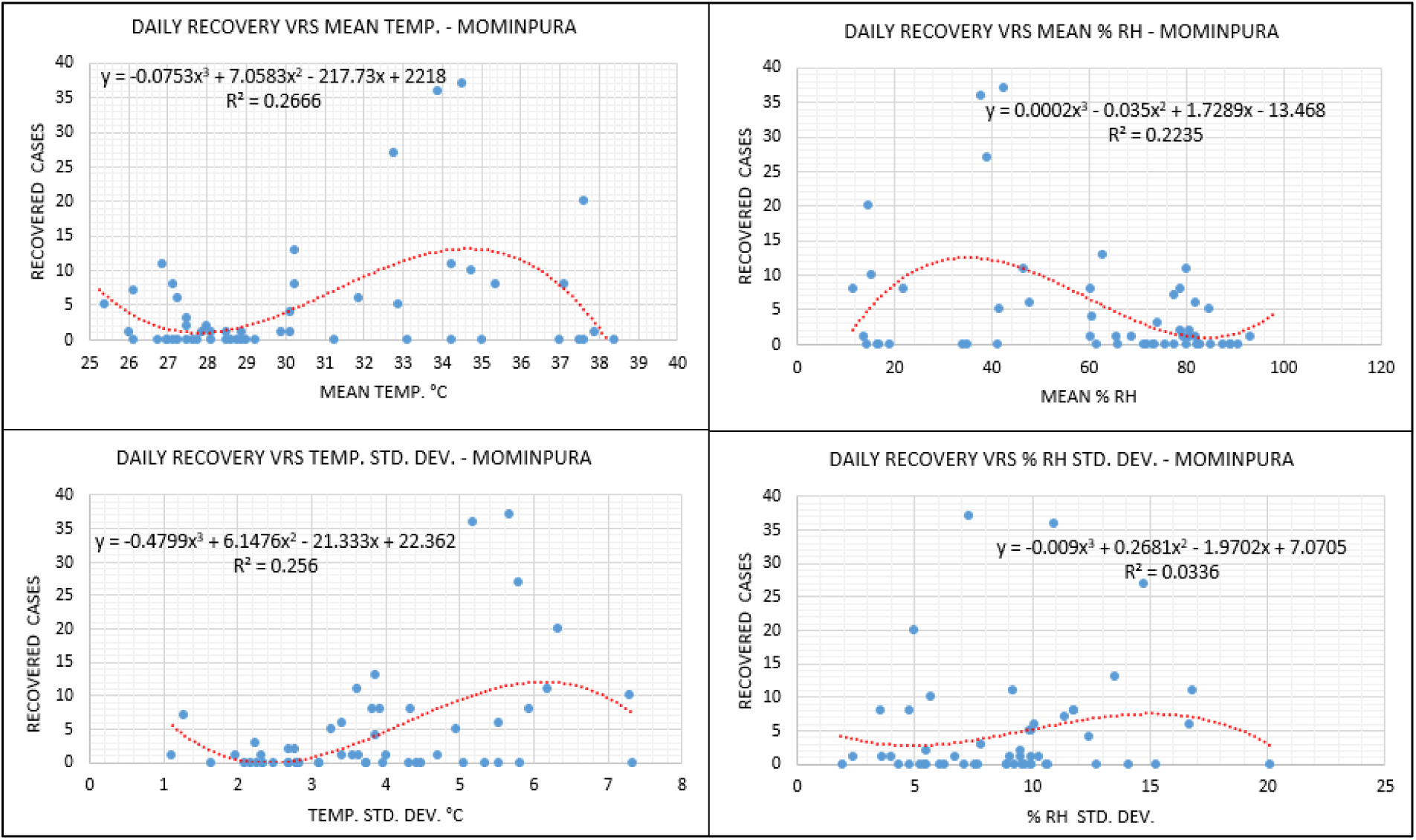
Environmental analysis – Mominpura (Nagpur MNC)

**Fig.19.**
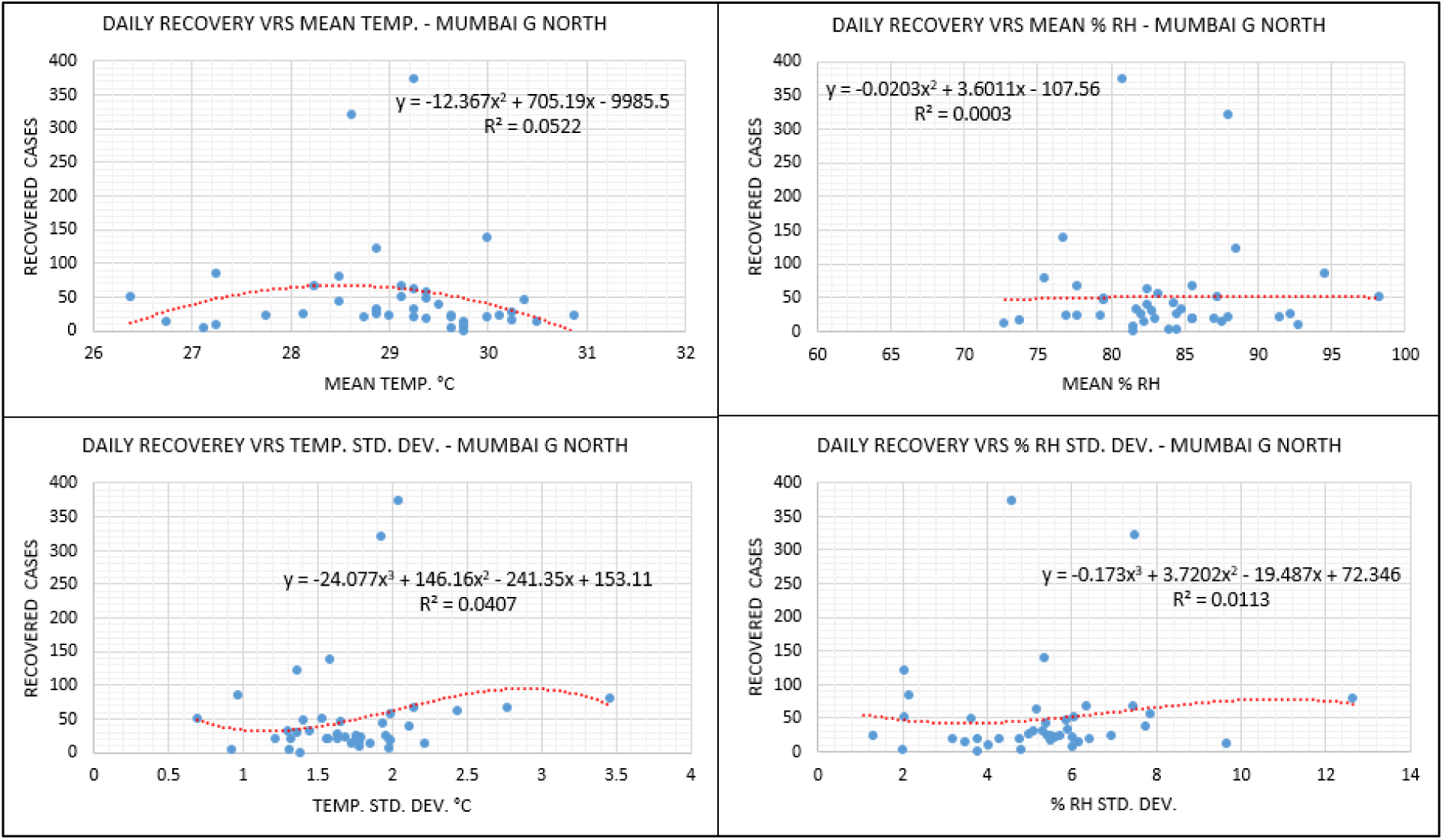
Environmental analysis – Mumbai GN ward.

**Fig.20.**
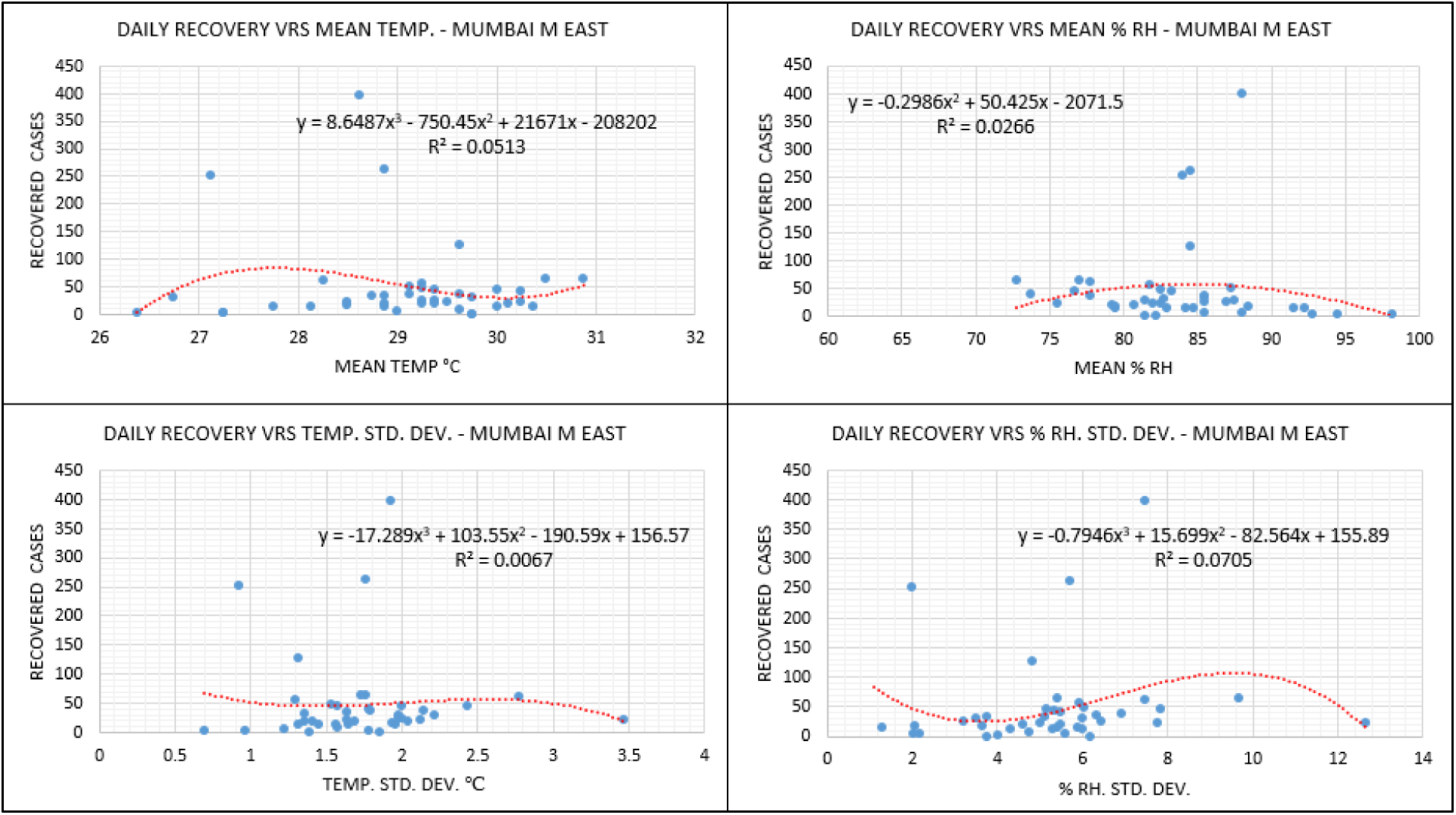
Environmental analysis – Mumbai ME ward.

**Fig.21.**
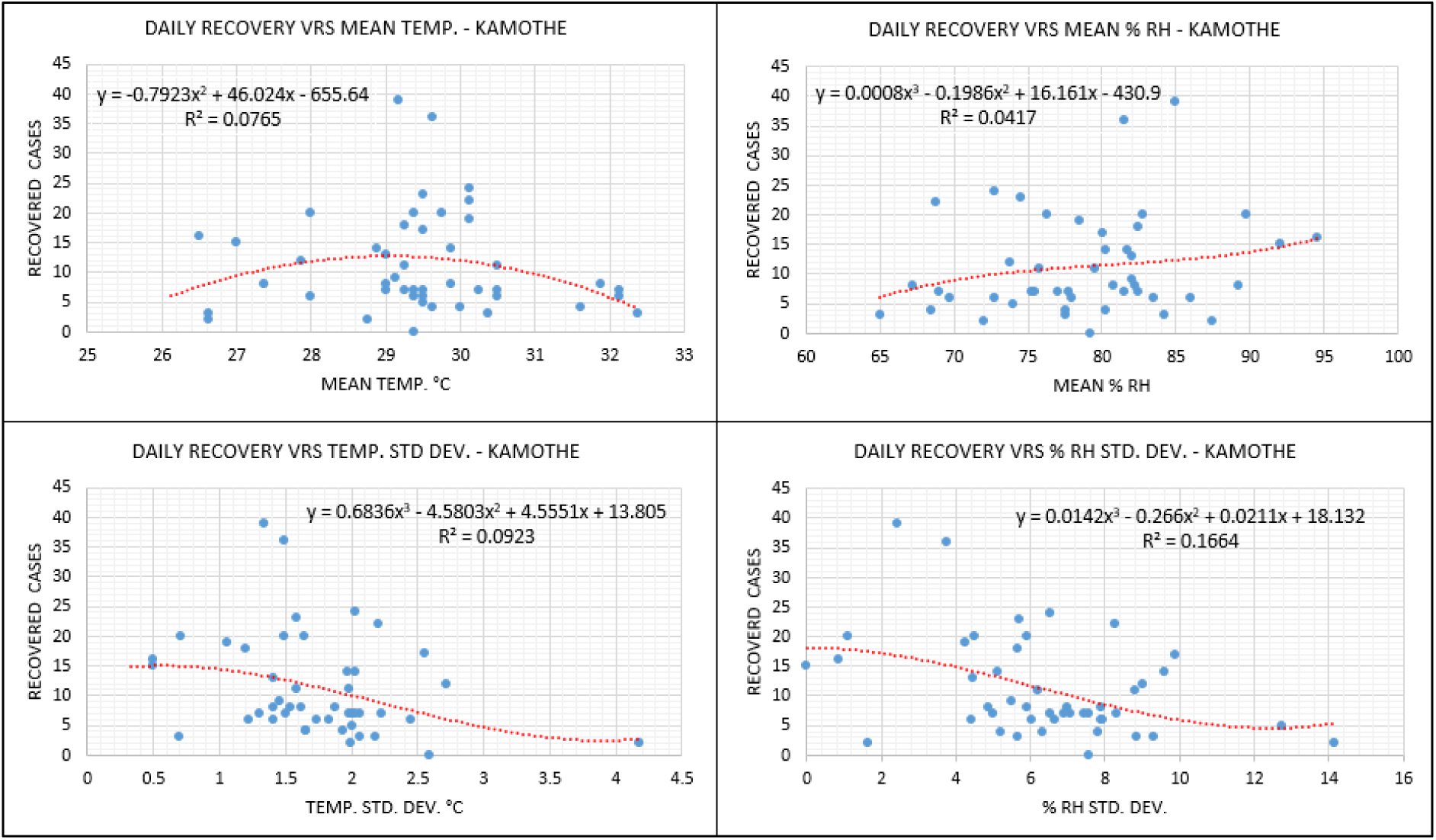
Environmental analysis – Kamothe (Panvel)

**Fig.22.**
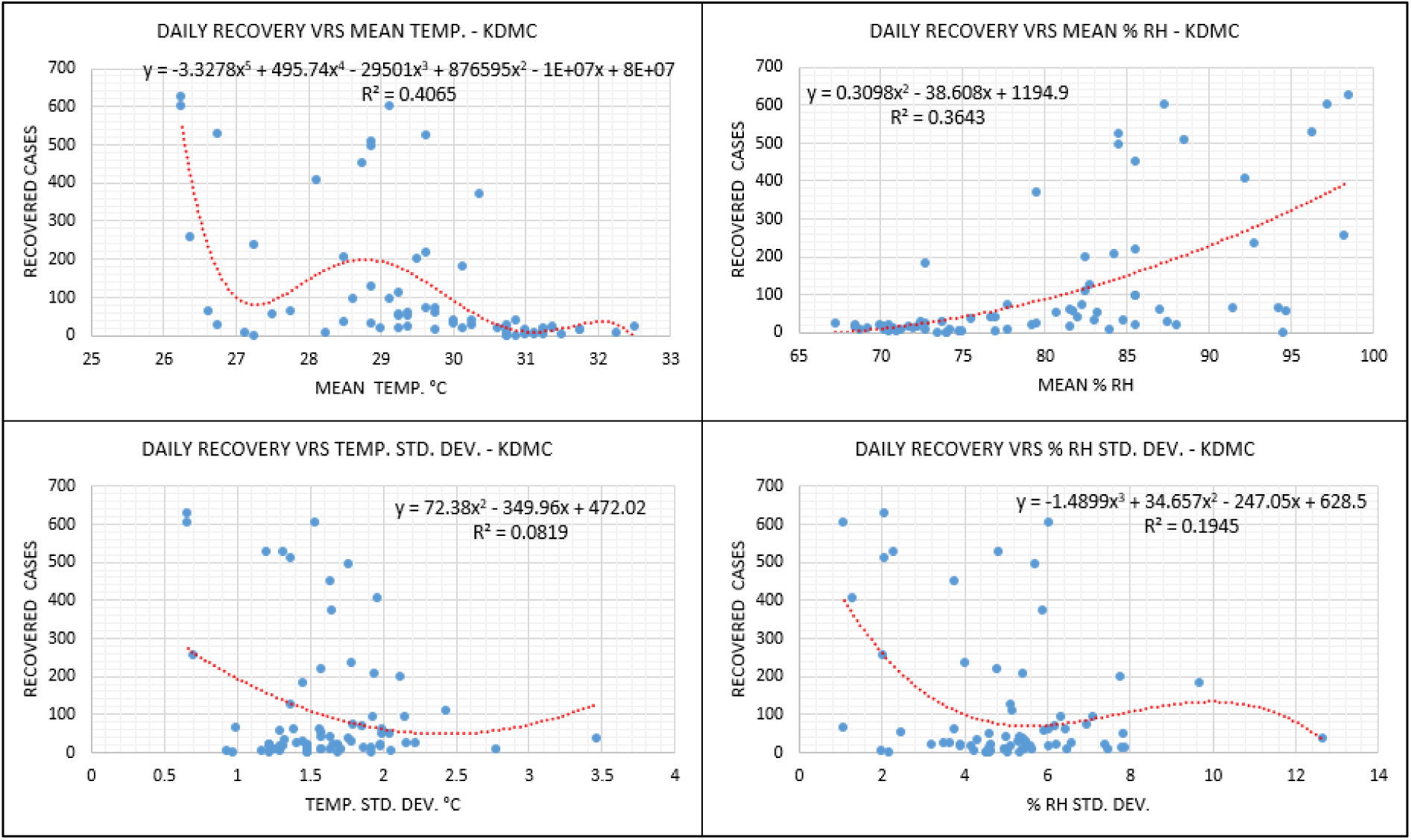
Environmental analysis – KDMC.

**Fig.23.**
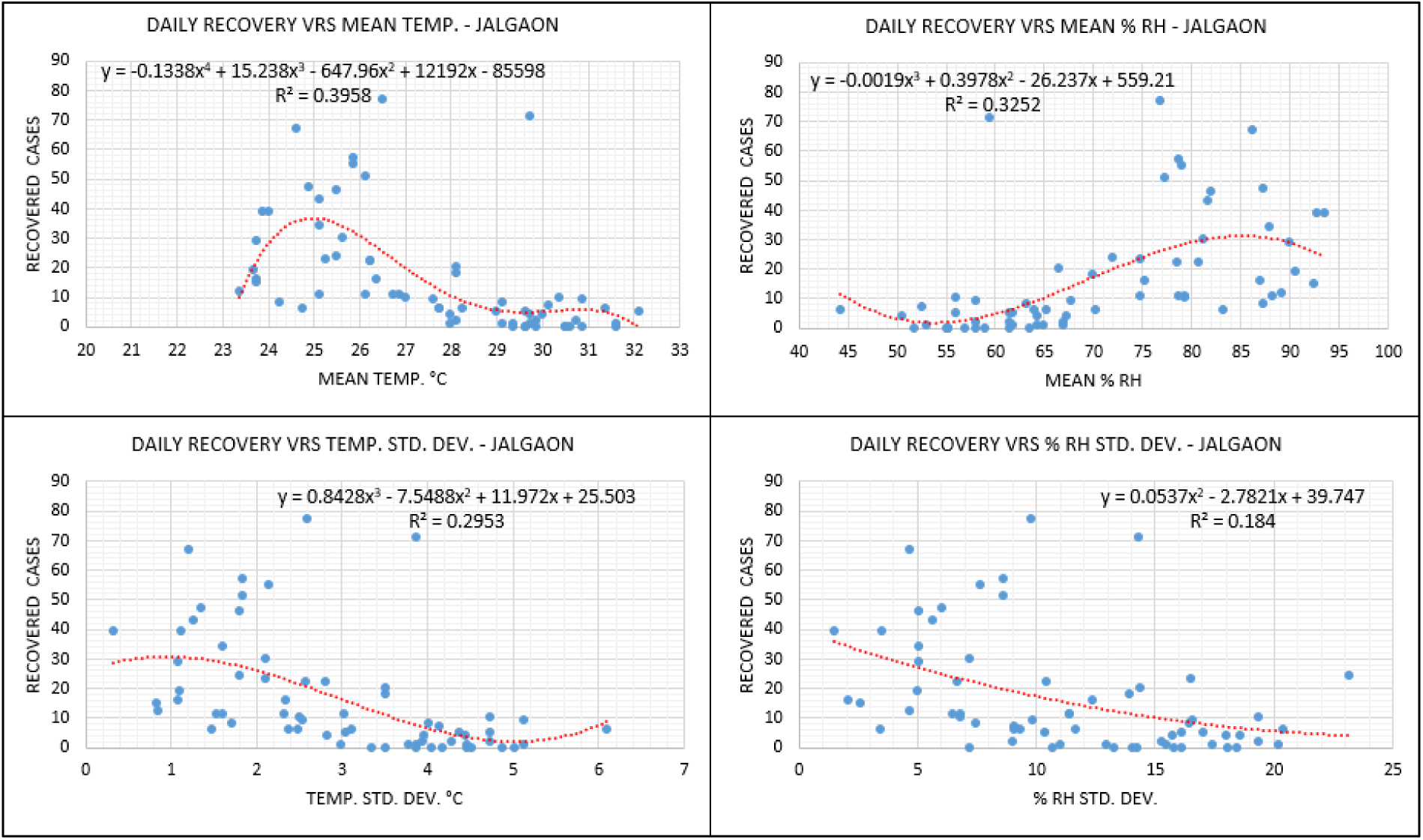
Environmental analysis – Jalgaon MNC.

**Fig.24.**
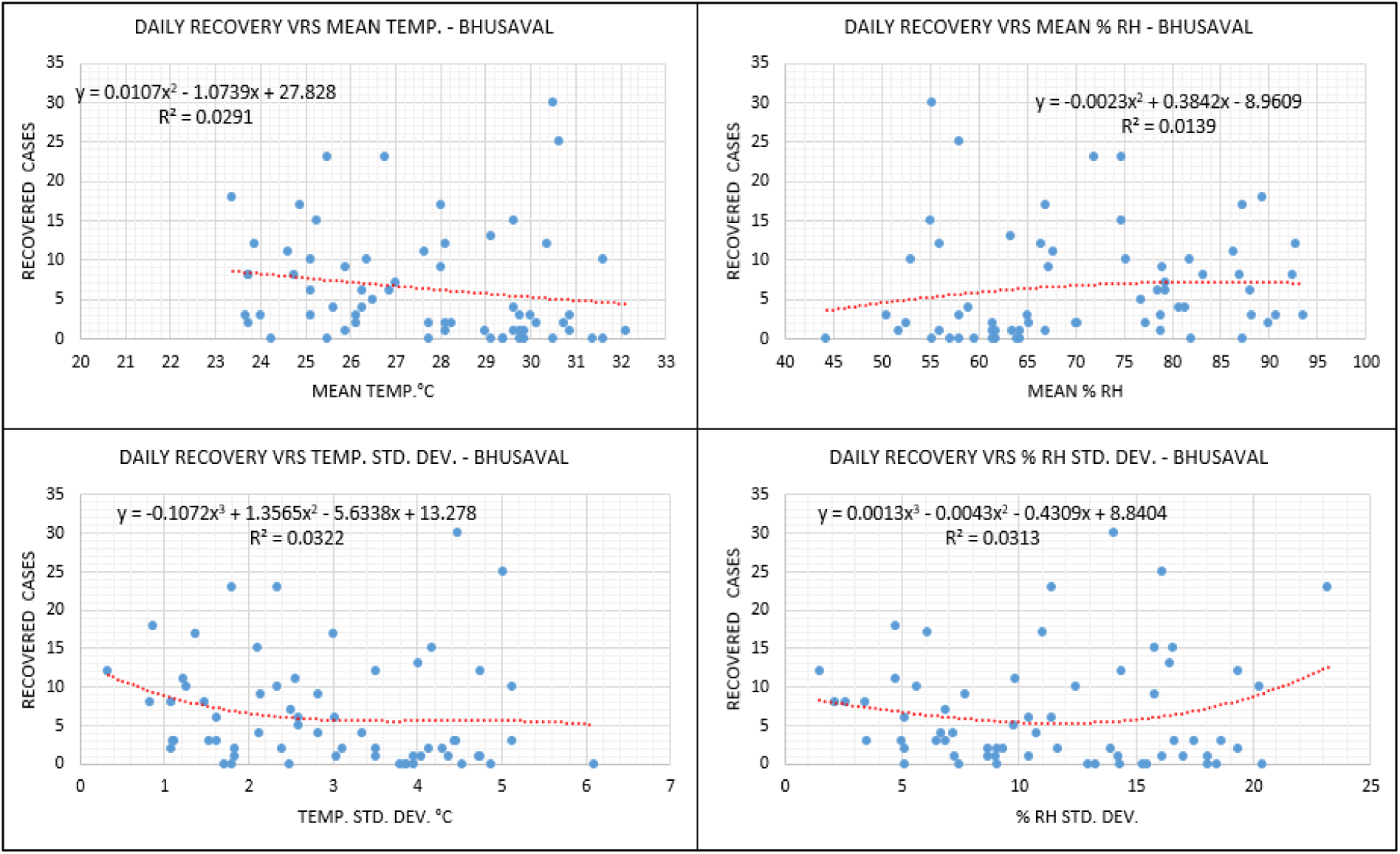
Environmental analysis – Bhusaval.

**Fig.25.**
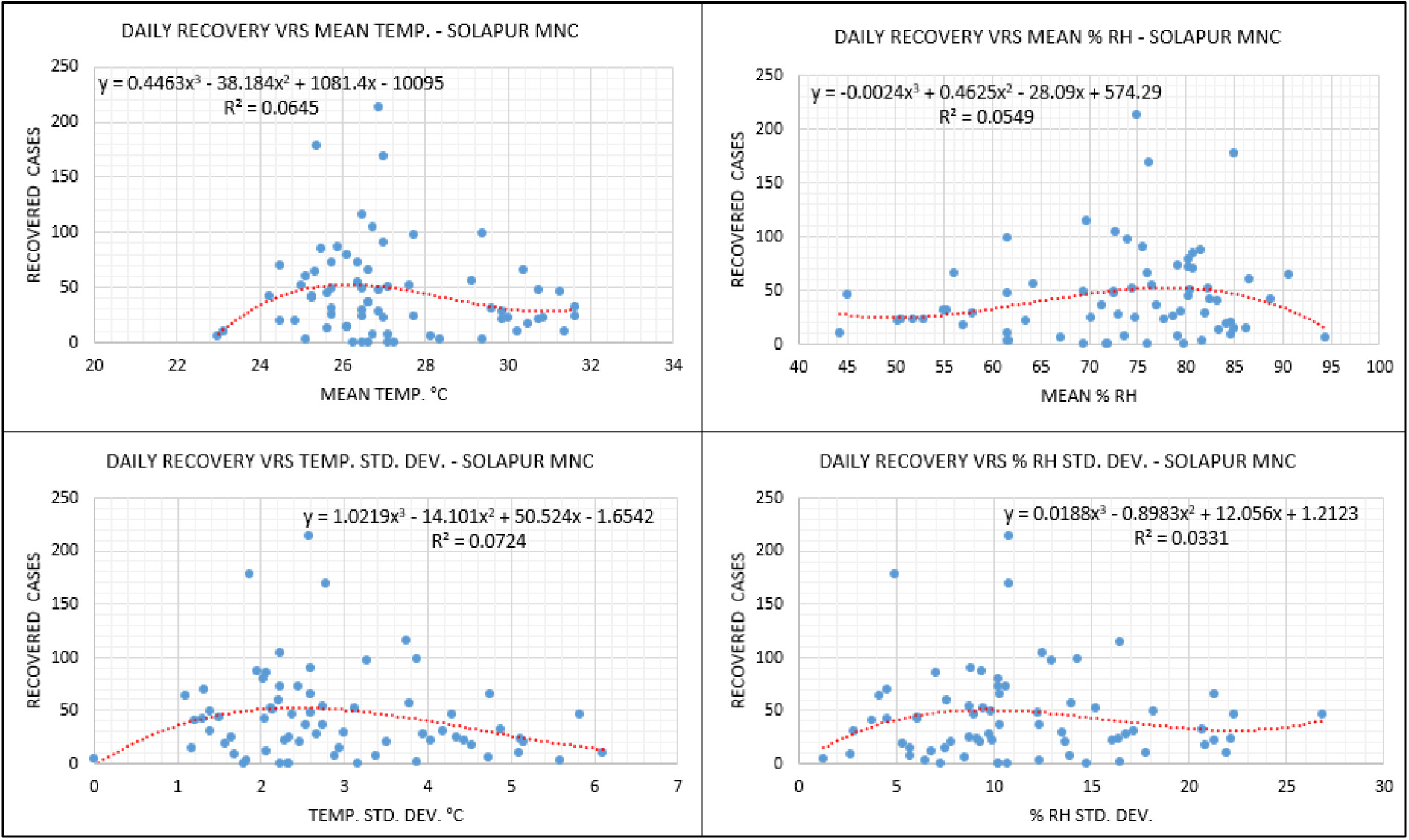
Environmental analysis – Solapur MNC.

**Fig.26.**
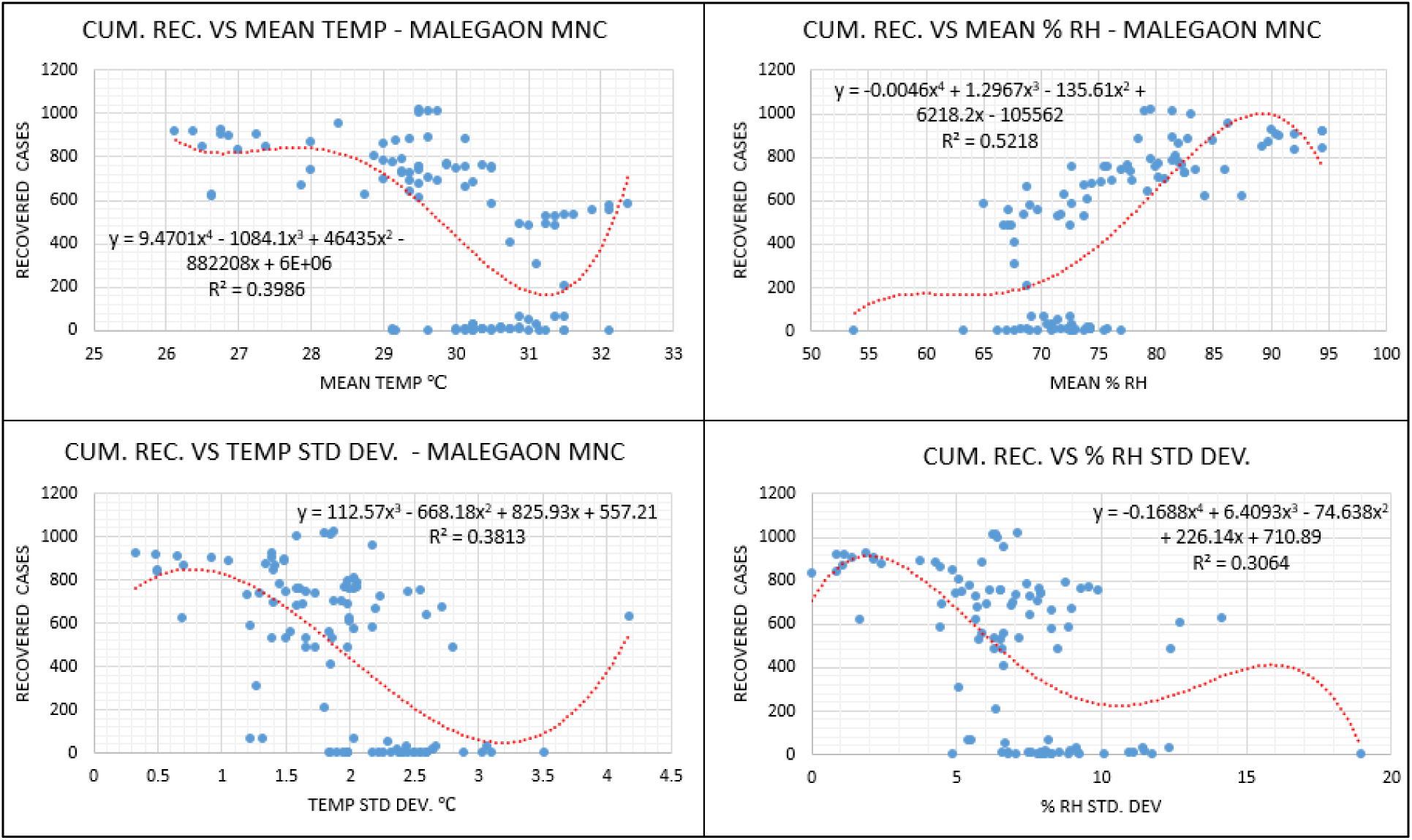
Environmental analysis (cum. data) – Malegaon MNC.

**Fig.27.**
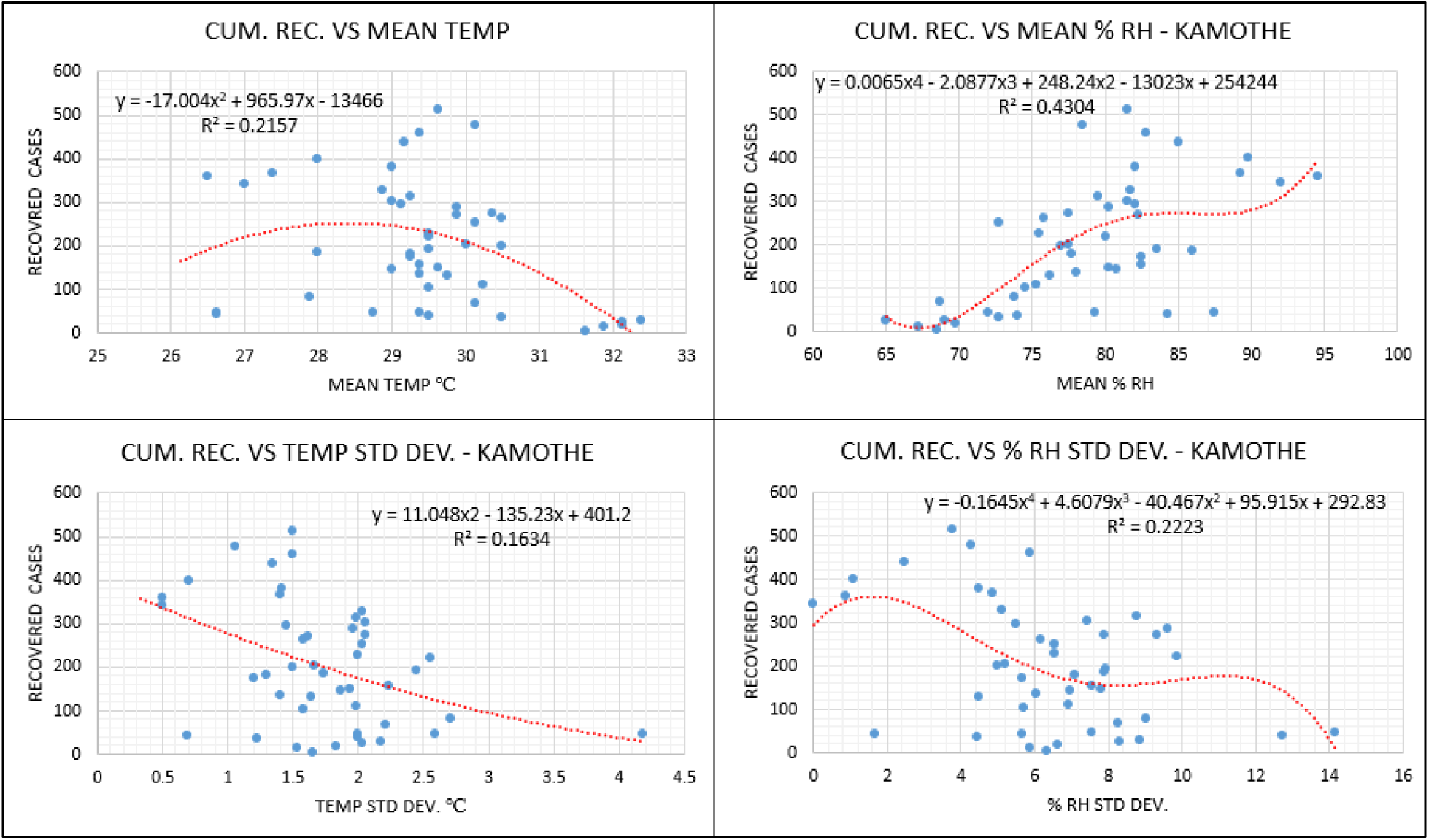
Environmental analysis (cum. data) – Kamothe.

**Fig.28.**
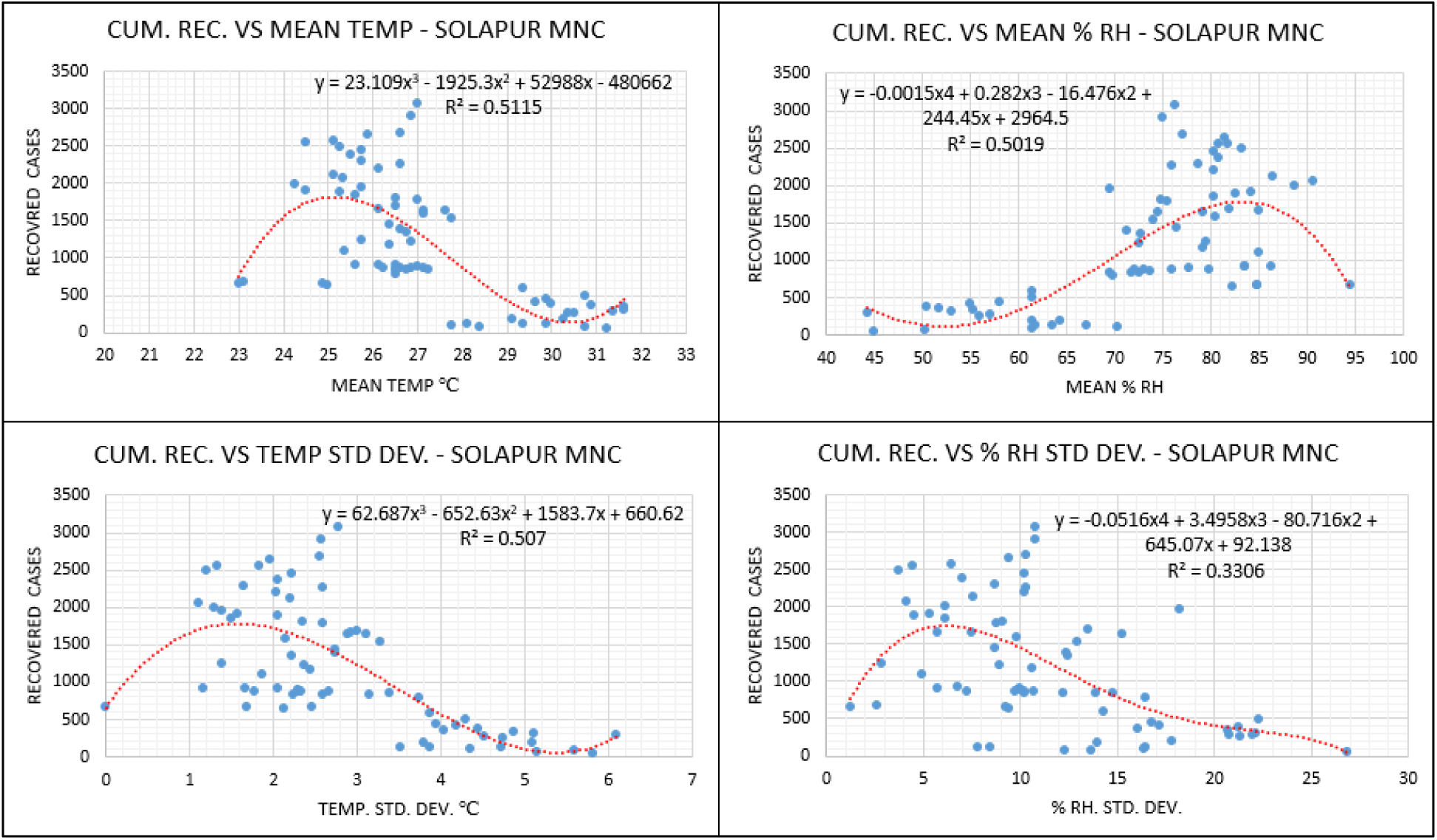
Environmental analysis (cum. data) – Solapur MNC.

R-square values are lower for Malegaon, Mumbai GN, Mumbai ME, Bhusaval and Solapur MNC whereas these are large for KDMC, Sataranjipura, Mominpura, Jalgaon MNC and Malegaon. For Kamothe, these values are lower as well except for active cases, where R-square is moderate as long as pandemic such as present one is considered, Fig.16-Fig.25. This is when data is analyzed on daily basis. On the other hand, when same data is plotted cumulatively, significant R-square values are noted. Of the three regions considered, Malegaon has highest R-square and Kamothe has least with Solapur in-between as far as recoveries are considered for mean RH, Fig.26 through Fig.28. Typical wavy nature can be seen through all the figures while navigating through scatters. Though the evaluation metrics in the case of daily data have values extremely low in the context of statistical significance, it is noticeable that daily recovered cases are increasing against the mean temperature range of 26° C to 30° C with deviations of less than 2°C (measured over 24 hours). Feasible range of mean relative humidity for the increasing number of the daily recovered cases are different for each containment zones depending upon geological positioning of the zone. However, it may be noted that daily recovered cases are increasing against a more dominant RH range of 70% to 85% and fluctuations of 1-15 % (for 24 hours). In our previous work [38], we have shown that higher relative humidity is responsible for an increase in the number of cases. However, it is essential to note that the same higher range of humidity is responsible for an increase in recovered cases.

Active cases as of July 31 2020, for all the containment zones under consideration, are analyzed against a feasible range of mean temperature, mean % RH, temperature deviations and % RH deviations in which significant amount of batch recoveries are observed. This is shown in Fig.29 (a) and Fig.29 (b). Fig.29 (b) is scaled version of Fig.29 (a) to have more insight in view of X-axis. As can be seen from the figures, wavy nature, typically of a sinusoidal kind with fluctuations in amplitude, is observed. In-fact such a nature is consistent for all the figures almost (Fig.16-Fig.30). This suggests that the active cases will fluctuate in the context of mean temperature and relative humidity with intermittent peaks of less amplitude than the first ones. This is, of course, possible if the undue disturbance of the public is not happening. Otherwise, a severe increase in active cases may happen. This is valid for Fig.16 through Fig.28. For brevity, other things of statistical variance and comments where significant lower R-square at different occasions are not detailed here. Nevertheless, it is understood that for the typical viral outbreaks like COVID-19, higher variances are expected.

**Fig.29 (a).**
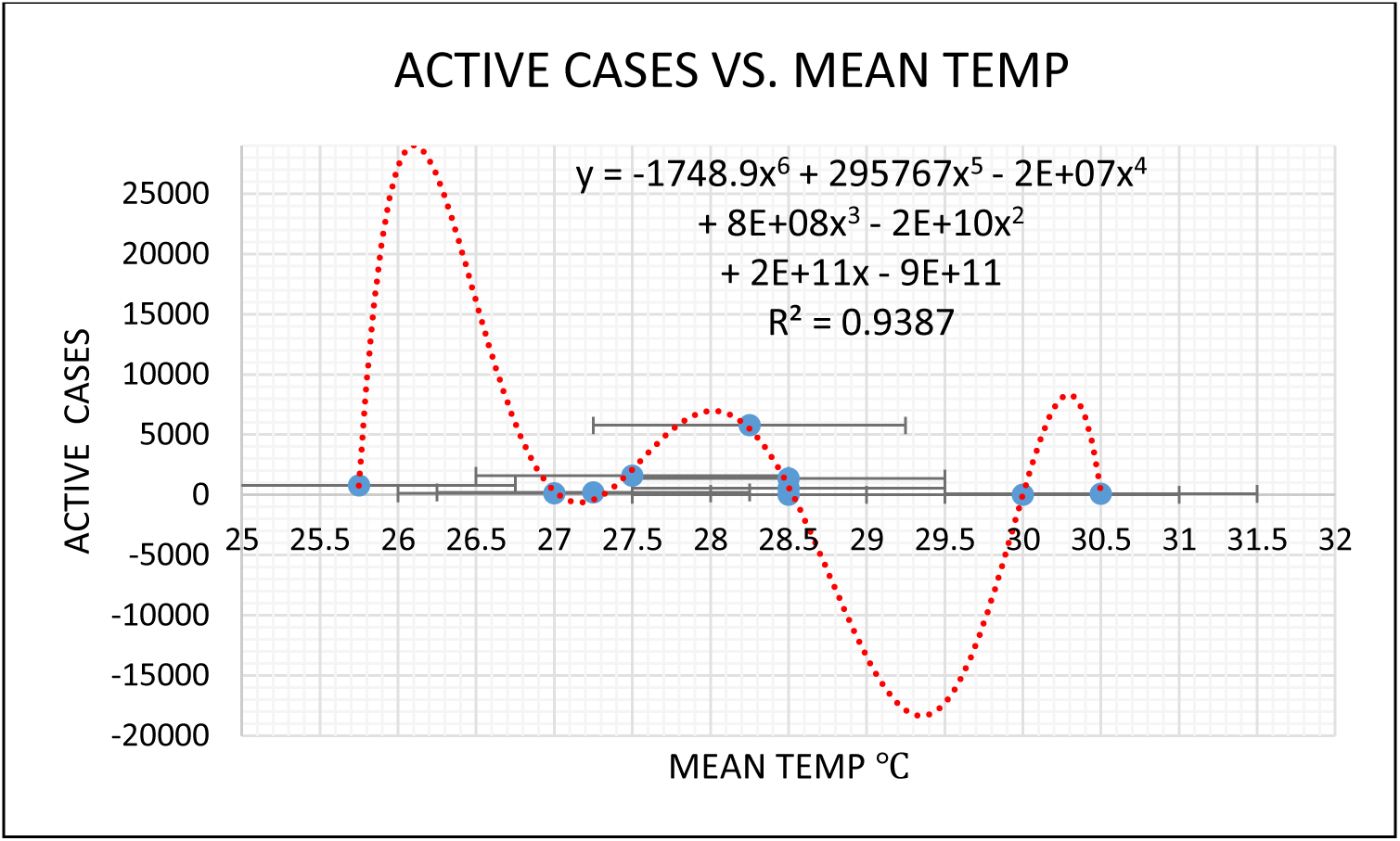
Active cases (July 31) vs feasible mean temp.

**Fig.29 (b).**
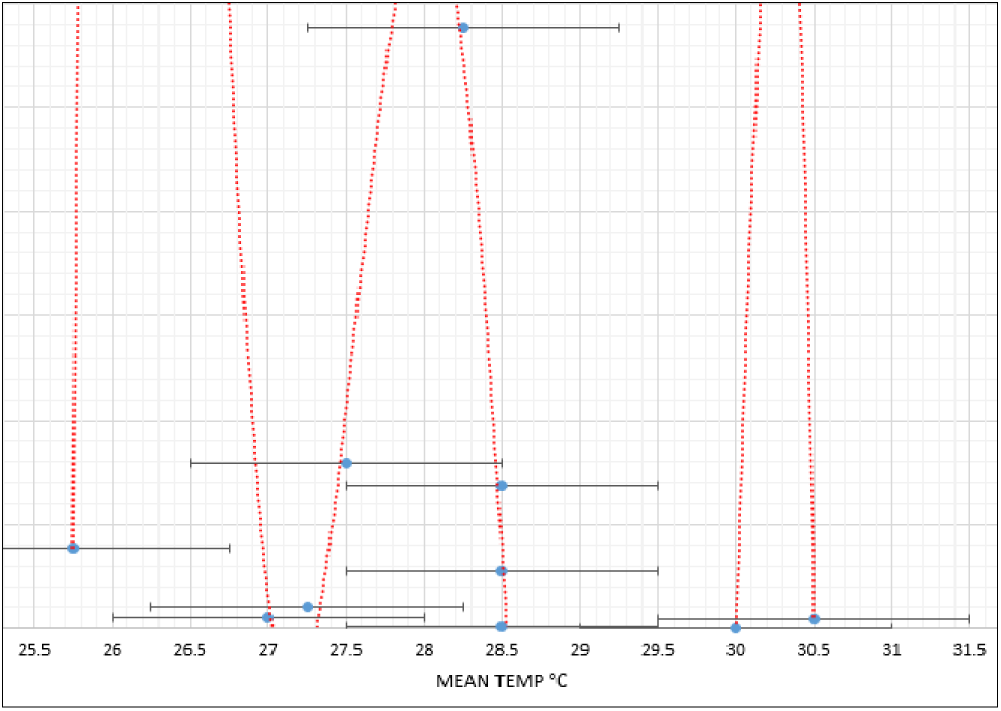
Active cases (July 31) vs feasible mean temp (scaled).

**Fig.30.**
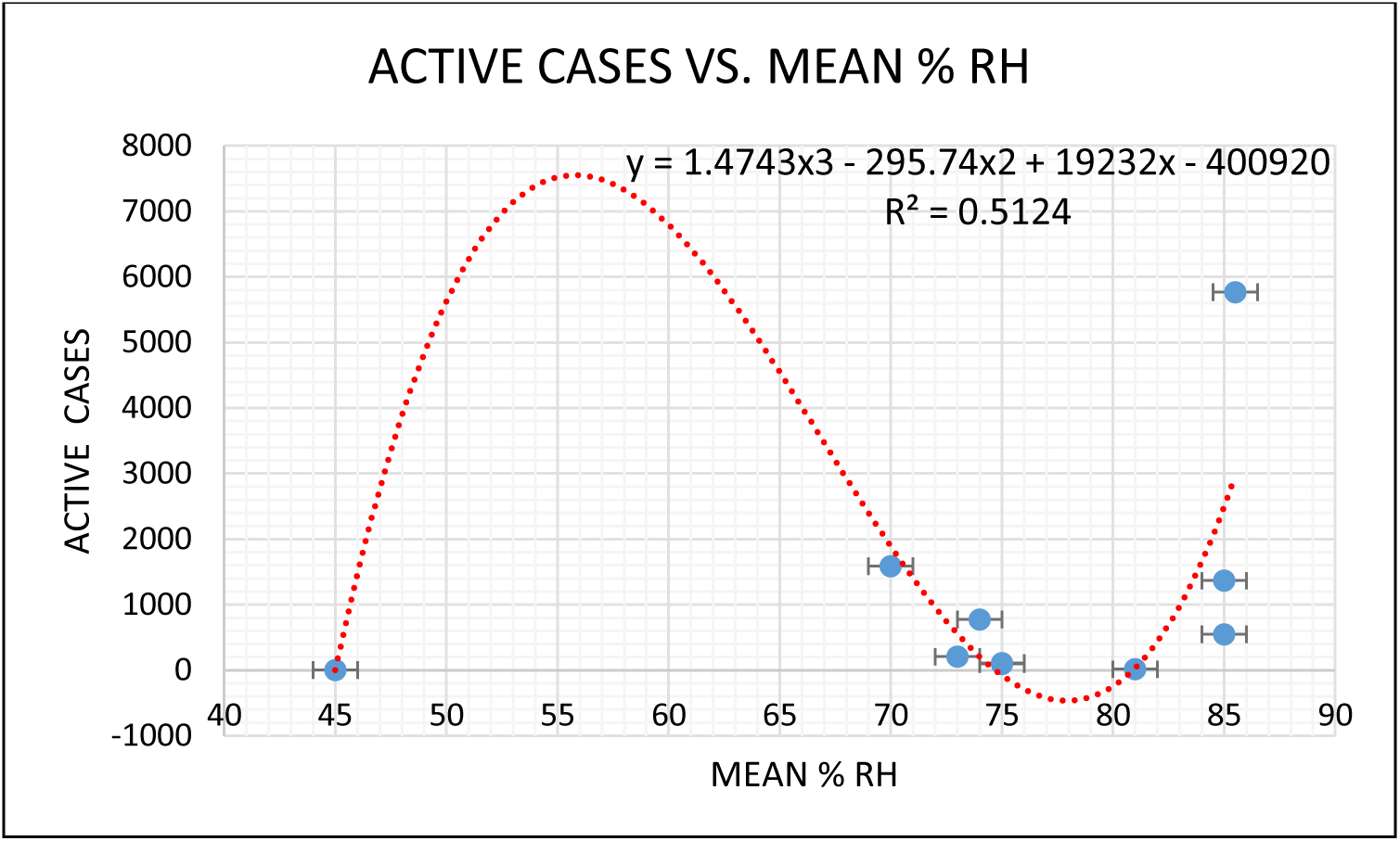
Active cases (July 31) vs feasible mean %RH.

### Behavioural factors

As per the earlier discussion, the environmental factors alone may not explain the outbreak pattern as well as the recovery scenario. The psychological behaviour of people in a particular community also plays a vital role in achieving of required outputs. Reduced earnings, loss of jobs, increase in unemployment, drastic changes in financial management etc. due to pandemic are some of the primary reasons behind increasing psychological stress in people of different communities. Immunity disorders are generally accompanied by individuals having higher stress levels. Among these disorders, the most damaging one is related to the cytokine storm.

**Table 2:** Cytokines and side effects (Cytokine storm) [39].

| **Cytokine** | **Producer Cell** | **Category** | **Neuropsychiatric side effects** |
| --- | --- | --- | --- |
| IFN-alpha | leukocytes | Pro-inflammatory | Fatigue, psychomotor slowing, depressed mood, anxiety, social withdrawal, anorexia, cognitive disturbances |
| IFN-beta | Fibroblasts | Pro-inflammatory | Fatigue, depressed mood, cognitive Impairment |
| IL-1 | Macrophages | Pro-inflammatory | Cognitive Impairment |
| IL-2 | T cells | Pro-inflammatory | Fatigue, anhedonia, dysphoria, cognitive Impairment ( mental slowing) |
| TNF-alpha | Macrophages, NK cells | Pro-inflammatory | Fatigue, Anorexia |

Abbreviations: NK, natural killer; TNF, tumor necrosis factor.

In Maharashtra, out of 4144 deaths whose comorbidity status is available are analyzed, it is found that comorbidities were present in 2898 cases, i.e., 70 % [36]. This suggests that individuals having existing multiple disorders and exposed to the COVID-19 virus may be considered as high-risk individuals in terms of possible fatality.

In the earlier period of this pandemic, the psychological stress and hence deaths due to comorbidities were limited to zones in Mumbai metropolitan regions. Gradually when lockdown restrictions were lifted, migration was started towards the other parts of the state. This resulted in an increase in the number of positive cases in other zones like Jalgaon MNC, Bhusaval, and Solapur MNC. Ultimately this created social havoc causing waves of social anxiety disorders and major depressive disorders in these areas. Though the other factors can also be vital, this might be the reason for the higher deaths in Solapur MNC where the drastic surge in active cases can be seen post-May 2020. In general, the rural population of the corresponding containment zones are under state of more fear, anxiety, stress, lack of awareness, old-traditions, too much casual nature and hypertensive behavior etc. this is responsible for rapid increase in the infected cases, reduced recovery and therefore more active cases.

### Virus mutation, recombination and social demography

Highly dense housing nature in some of the zones under consideration makes it almost impossible to avoid contact within the residents of the community. This might eventually lead to infecting around 50 to 60 percent of people of the total population; as a result, the mutation of the virus takes place through genetic selection. In due course of infecting a large number of people, error may be incorporated in the genome of the virus. These mutations can be detrimental, neutralized, or sometimes favourable for the community. If the essential virus functions are interfered with or interrupted during these mutations, the further spread may not be persisted in such a community. See Chapter 43-Viral Genetics [40]. However, this process may take an adverse toll on individuals with high levels of stress by means of hypertension, fear as well as anxiety.

In socially isolated communities like Sataranjipura, Mominpura, and Malegaon MNC controlled mutations might alter the viral strain in the context of the airborne range up to which the host can get infected. It is also possible that virus strength had reduced for specific cell target mechanism. The overall effect may be expected as mutations leading to reduced pathogenicity of virus strain keeping its antigenicity intact. Such type of virus mutations can be used to extract vaccine strains. Hence it can be concluded that the possibility of herd immunity can be highly expected in such socially isolated communities. On the other hand, in the Mumbai metropolitan region, fundamental norms of socially isolated communities are challenging to follow. Also, the recovery of cases, as well as the reduction in active cases, are much slower than expected due to the mixed population. The main reason behind this is the interdependency of different areas in daily life in the Mumbai metropolitan region. There is a continuous movement of passengers and goods on a daily basis in the context of essential services over the wider metropolitan area of 6355 km2. Owing to this reason, there are phenotypic variation occurrences in the present strain of novel coronavirus. The uncontrolled mutations might lead to producing antigenic drifts, which might result in a modified strain of virus capable of attacking the hosts, those are resistant to earlier strain. Further, there is also a possibility of recombination of the present virus strain with similar viruses in the process of coinfection of a host cell. This is due to the exchange of genetic materials along with virus spread occurring due to the high volume of public interactions over broad areas of the metropolitan region.

In a nutshell, regions like Mumbai, Kamothe, KDMC, and Jalgaon MNC have a significant mixed population. Also, community isolation is practically not possible in these regions, as discussed above. Due to this, mutation and recombination of virus strain are higher in due course of infecting a large number of people. As a result, the infection spread in these regions is of rapid increasing nature. On the other hand, containment zones like Malegaon MNC, Sataranjipura, Mominpura and Bhusaval have a dominant particular community population.

Moreover, these containment zones have shown excellent isolated community behaviour throughout the lockdown period. This spread of infection has been faster, resulting in early saturation of viral infection. The size of the virus in high humidity areas is larger as compared to areas having less humidity. Further due to larger aerosol size in coastal regions (due to high humidity) than the non-coastal regions, asymptomatic cases are more likely in the former region whereas more symptomatic cases will result in the latter region. These effects will have severe consequences on COVID-19 outcomes if medications are inadequate in non-coastal regions.

### Diet and social demography

Another most important factor which plays a crucial role in maintaining a good immunity system is diet. Maharashtra is well known for the inclusion of a variety of spices and fried food items in the diet. The imposition of lockdown since March 22, 2020, has created much havoc across the state and forced people to remain at home. This resulted in an abrupt change in the lifestyle of most of the people and halted their habits of various kinds of outdoor exercises through walking, running, swimming, gyms etc. At the same time, most of the people in urban areas like Mumbai, Thane, and Nagpur preferred to eat a variety of spicy and fried types of food items. This increased laziness, cholesterol, obesity problems. After the lockdown period, the average consumption of caffeine by individuals has increased due to the consumption of tea and coffee considered as a hot beverage which is remedial solution. Due to this increased amount of caffeine consumption, many people started facing anxiety problems as caffeine is a central nervous system stimulant that increases anxious feelings [41]. In the earlier period of lockdown, containment zones like Mumbai GN, Mumbai ME, KDMC, and Kamothe had faced a shortage of vegetable supplies due to restrictions on movement. Lack of a balanced diet also resulted in vitamin and other deficiencies in the majority of individuals in urban areas across the state. Following Table 3 shows essential food contents, its sources, health benefits, results of deficiencies and regions in which particular food constituent is dominantly consumed through typical dietary patterns.

**Table 3:** Food data Chart.

| Essential Food constituents | Sources | Health benefits | Deficiency results | Regions enriched |
| --- | --- | --- | --- | --- |
| VITAMIN A | Indian salmon, Tuna, Surmai fish, Eggs, Carrot, Sweet Potato, broccoli, Green leafy vegetables, milk, yoghurt and other dairy products. | • Boost immune function<br>• Maintains the heart, kidneys and lungs function.<br>• Moistening mucous membranes.<br>• Promoting cell growth. | • Nerve damage<br>• Reduces resistance to respiratory illness.<br>• Dry mucous membrane. | • Mumbai GN and Mumbai ME<br>• Kamothe<br>• KDMC |
| VITAMIN B-12 | Found in animal products including fish, meat, chicken, eggs, milk and milk products, fortified cereals. | • Necessary for the production of mood chemicals in the brain.<br>• Reduces risk of cardiovascular diseases. | • Fatigue, weakness, loss of appetite, weight loss.<br>• Difficulty in maintaining balance.<br>• Depression.<br>• Confusion. | • Mumbai GN and Mumbai ME<br>• KDMC<br>• Sataranjipura<br>• Mominpura<br>• Malegaon<br>• Solapur MNC |
| VITAMIN C | Oranges, lemons, strawberry, Green leafy vegetables, green and red peppers, kiwi, tomato juice, broccoli. | • Strengthens body's natural defense.<br>• Boost the immune system.<br>• It may help to manage blood pressure.<br>• It may lower the risk of heart diseases.<br>• Good for physical and mental health<br>• Improves absorption of iron from diet. | • Increases the chances of developing viral flu.<br>• Sufferings from cold.<br>• Joint and muscle pain<br>• Tiredness and weakness | • Sataranjipura<br>• Mominpura<br>• Mumbai GN and Mumbai ME<br>• KDMC<br>• Kamothe |
| VITAMIN D | Sunlight exposure, fish, egg yolk, orange juice, mushrooms, COD liver oil, milk and milk products. | • Reduces the risk of developing several chronic diseases.<br>• protects respiratory tract<br>• kills enveloped viruses<br>• decreases production of pro-inflammatory cytokines<br>• Plays key roles in brain and nervous<br>system, helping to maintain normal function. | • Development of autoimmune diseases.<br>• Increases susceptibility to developing illnesses and diseases<br>• Fatigue and tiredness<br>• Depression | • Mumbai GN and Mumbai ME<br>• KDMC<br>• Sataranjipura<br>• Mominpura<br>• Malegaon<br>• Solapur MNC<br>• Jalgaon MNC<br>• Bhusaval |
| OMEGA 3<br>AND OMEGA<br>6 | Fish and various seafood, soybean, cod liver oil, sunflower oil, corn, meat, eggs. | Cell growth, brain development, and nerve message transmission.<br>Maintains a healthy heart, ameliorating inflammatory diseases, reducing bad cholesterol, promotes brain development. | Risk of depression and anxiety disorders.<br>Risk of heart attack<br>Risk of blood clotting<br>Higher susceptibility to infection.<br>Bad memory performance | Mumbai GN and Mumbai ME<br>KDMC<br>Sataranjipura<br>Mominpura<br>Malegaon MNC |
| IRON | Meat, chicken, eggs, seafood, beans, leafy vegetables | Boosts hemoglobin, which helps to transport oxygen in the blood.<br>It may help manage unexplained fatigue.<br>Strengthening immune system.<br>Improves muscle endurance. | Unusual tiredness<br>Shortness of breath<br>Headaches and Dizziness<br>Chest pain, fast heartbeat | Mumbai GN and Mumbai ME<br>KDMC<br>Jalgaon MNC<br>Bhusval<br>Solapur MNC |
| ZINC | Fish and various seafood, chicken, meat, eggs, beans, milk and milk products, vegetables. | It helps keep immune system healthy.<br>May reduce risk of age-related diseases<br>Zinc decreases oxidative stress, which may lead to chronic inflammation. | Major depression<br>Risk of diabetes<br>loss of appetite<br>chronic kidney failure | Mumbai GN and Mumbai ME<br>KDMC<br>Solapur MNC<br>Kamothe |

Due to lockdown restrictions, it is believed that the dietary pattern of the majority of people across the state has been disturbed and changed. Not maintaining the minimum amount of food constituents in the routine diet may lead to an increase in comorbidities, which can be an alarming situation in the context of the COVID-19 viral outbreak and post COVID-19.

The following Table 4 gives an overview of demography, general appetite in containment zones, and problems that occurred in the context of diet during the lockdown.

**Table 4:** Social demography and diet.

| SR. NO. | Areas | Routine Dominant Food Content | General Issues during Lockdown |
| --- | --- | --- | --- |
| 1 | Mumbai Wards: GN and ME | Variety of vegetables and fruits<br>Fish, Meat, Rice items, Fast food and processed food. | Unavailability of vegetables as well as Meat and fish during earlier period of lockdown. |
|  | • KDMC<br>• Kamothe |  | • Fear of covid-19 spread through meat caused many people to avoid it.<br>• Increase in caffeine intake. |
| 2 | • Jalgaon MNC<br>• Solapur MNC | Locally grown fresh Vegetables and fruits, sorghum, millet etc. | • Unavailability of vegetables and fruits in containment duration.<br>• Fight shy of meat in diet.<br>• Overall, significant population avoids onion, garlic and traditional spices due to community beliefs. |
| 3 | • Malegaon<br>• Sataranjipura<br>• Mominpura<br>• Bhusaval | Variety of meat products presiding in diet, vegetables and fruits in adequate quantity. Inclusion of variety of mixed spices in daily diet | • Unavailability of vegetables in lockdown period, driven majority of people to higher consumption of Meat and related food items. |

In everyday cooking of vegetables and non-veg items across the state, spices play a crucial role in how the different communities cook and consume food. Traditional Indian spices made from a variety of ingredients have several health benefits, as shown in the following Table 5.

**Table 5:**
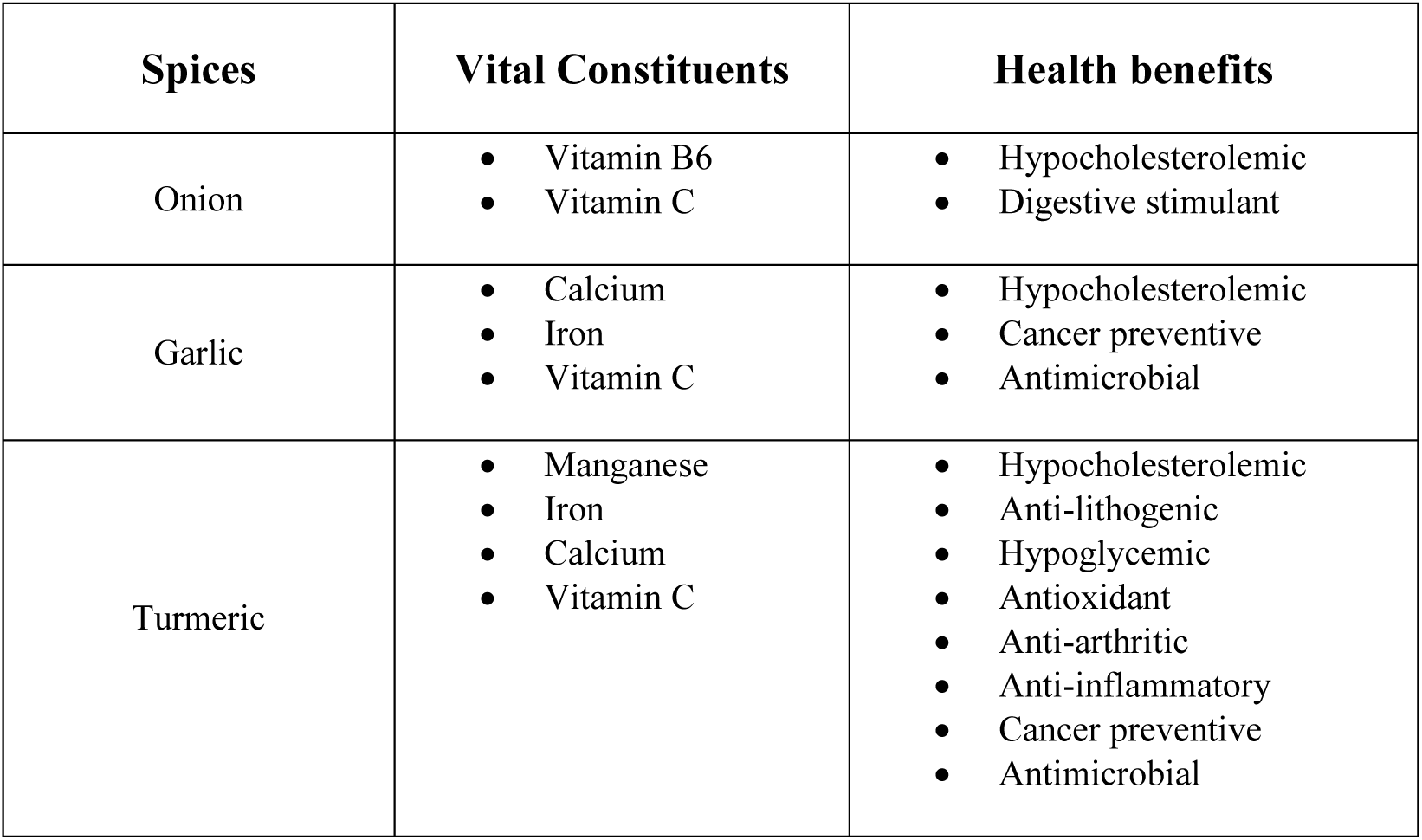
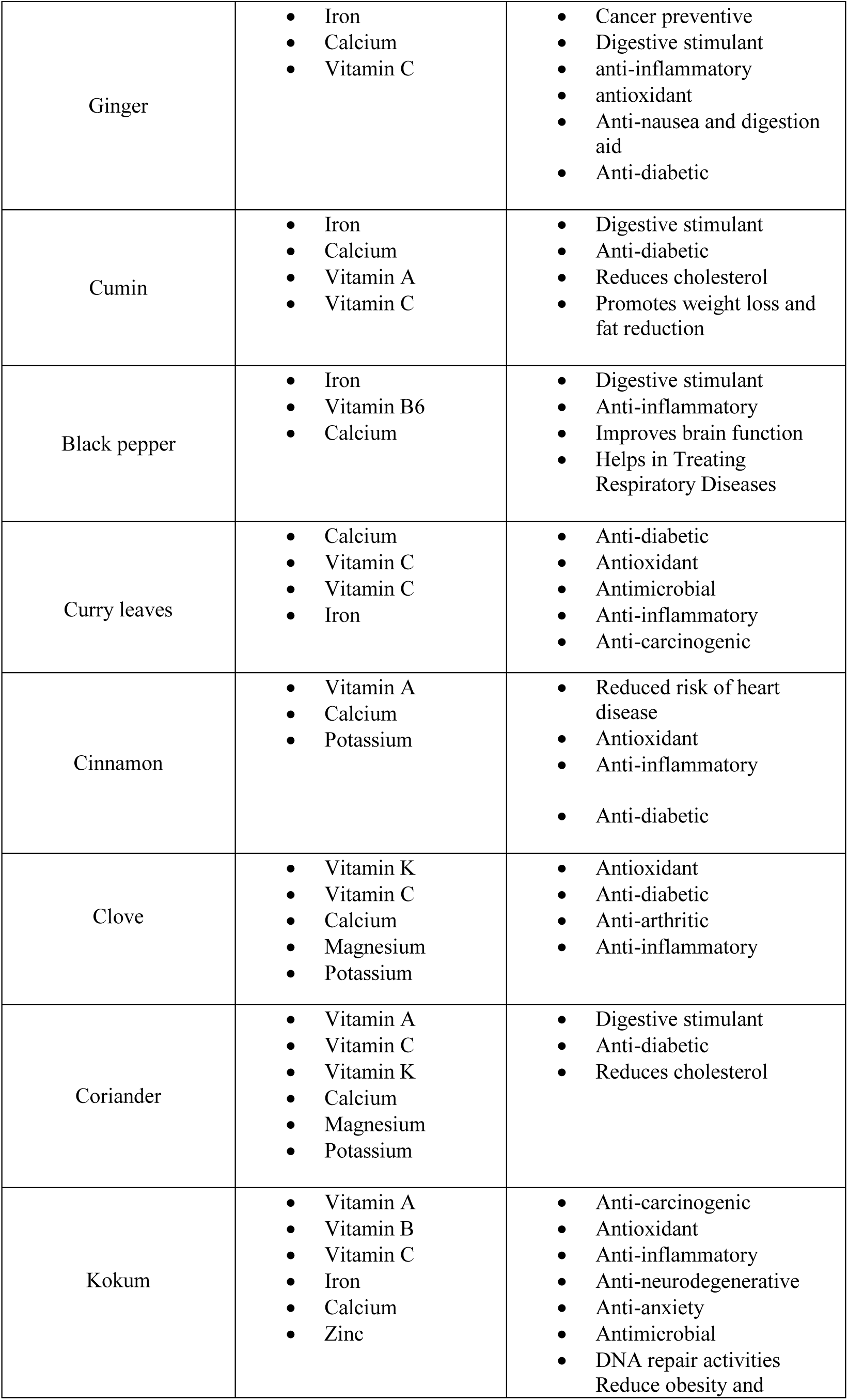
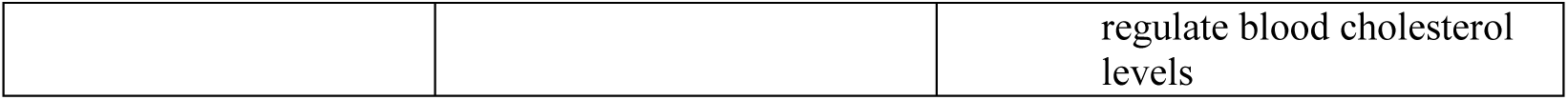
Constituents of Maharashtrian spices.

It may be possible that due to the lockdown restrictions (Unavailability of vegetables, meat, and fish for some period) that the routine consumption of these spices has been disturbed in the diet of many people across the state. The deficiency of essential constituents, as mentioned in the above table, might also lead to an increase in several health issues, eventually contributing to the more comorbidities during the COVID-19 outbreak. Due to inherent dietary patterns, vegetarian and non-vegetarian diet, consisting the spices illustrated in Table. 5, have phenomenal significant positive effects on infected cases, recovered cases, deaths and thereby active cases as compared to the western countries. Still COVID-19 has changed us, in particular in the view of immunity. As outbreak is still existing, immunity of population is not as was before. Therefore, even weak strain of the virus become detrimental and has controlled or uncontrolled mutations.

### Mortality

While evaluating the impact of any viral outbreak, it is crucial to consider the deaths reported and the factors affecting it. Key factors that govern the fatality of novel coronavirus are lockdown implementations, isolated communities, diet, psychological and physiological behaviours, comorbidities, social demography, and availability of high tech medical care units for critical patients etc. Among all these factors, lockdown implementations played a crucial role in affecting the deceased cases in the initial days of the virus outbreak. Environmental factors like extreme humidity and temperature conditions, too, affect the pathogenicity of the virus and, eventually, the immunity of individuals in corresponding areas. Individuals with affected diet routines are suffering from immunity disorders and are at high risk of getting infected to the novel coronavirus. There are high possibilities of fatality in COVID-19 positive cases that have several medical conditions, i.e., comorbidities like heart, kidneys, respiratory issues, blood pressure problems etc. The significant impacts of this viral outbreak also resulted in economic losses, unemployment, financial insecurities etc. This has increased psychological stress in different communities at a higher level. Individuals with severe amounts of stress levels are highly likely to be turned into critical COVID-19 cases, which may lead to fatality.

As of July 31 2020, Solapur MNC had reported the highest mortality with 351 against 4985 total number of confirmed positive cases. It is also noticeable that Solapur MNC has a meagre recovery rate of 61 % among all the zones taken under consideration. Mumbai GN ward, Mumbai ME ward and Malegaon MNC containment zones have a high fatalities with 439, 292 and 85 deceased cases respectively, but it is essential to consider that population densities for the wards in Mumbai are 25358 / sq.km and for Malegaon MNC it is around 19000 / sq. Km. For Solapur MNC, which is having a population density of 5345 / sq.km, 351 deceased cases are very much higher. This is due to the possibility of a lack of extreme critical situation medical care units available in semi-urban parts like Solapur MNC. Also, Solapur MNC has a population scenario in which the majority of people in the different communities prefer to avoid consumption of non-veg food, traditional Indian spices that contain onion, garlic etc.

Containment zones like Mumbai ME and Mumbai GN, KDMC, and Jalgaon MNC, along with Kamothe in the Panvel MNC area, have a mixed population which highly prefer a variety of traditional Indian spices along with meat and seafood in the diet. But in these zones, the factors that drive the deaths are the high population density and, eventually, no practical social distancing in everyday routine. Also, due to the work nature, mutation and recombination of virus strains are very rapid, as discussed earlier in the MMR. This may cause severity in some COVID-19 positive cases having comorbidities leading to fatality. It is also important to note that some individuals with different community beliefs prefer not to take the diet that the majority of people follow in these areas. Such people could become high-risk individuals for getting infected and turning into severe COVID-19 cases. This is mostly due to their body’s inability to interrupt essential virus functions in the course of virus mutations or recombination. It may also result in failure in the development of antibodies due to very much different food content than the other majority of people.

These interrelated issues discussed above are believed to be tackled through some encouraging results seen in the containment zones of Sataranjipura and Mominpura of Nagpur MNC as well as Malegaon MNC. The deceased cases in Sataranjipura and Mominpura are 8 and 5 respectively as of July end, with the recovery rates of 85.77 % and 95.65 %. In the case of Malegaon MNC, though the deceased cases are relatively higher, the nature of the curve of the same is gradually decreasing. The reason behind the higher deaths in Malegaon MNC may be the increase in stress levels due to financial issues, occupational practices, ignoring early symptoms, hesitation to medication, hurdles in intense COVID-19 testing due to densely populated areas etc. Nevertheless, it is crucial to consider the significant recovered cases in Malegaon MNC with 87 % and 90 total active COVID-19 cases as of July end. This is due to fact that tracing is almost impossible in such areas and therefore it is more likely that more number of asymptomatic or symptomatic cases will be there unattended. Hence overall control of the viral outbreaks in Malegaon MNC and Nagpur MNC zones is entirely satisfactory. The main reason behind promising results emerging out of these three profoundly affected zones can be considered as dominant and isolated communities. Once put into the strict restrictions, Sataranjipura, Mominpura, and Malegaon MNC behaved as totally isolated zones over broad areas owing to very negligible in and out movements. As a result of community isolations and practical difficulties in maintaining social distancing practices due to dense population, a saturation of viral infections may have been reached, leading to a significant reduction of total active cases in a shorter period.

Jalgaon MNC and Bhusaval containment zones in the northern part of the state have scorching and dry weather conditions. Though metrological factors can be considered the same for both the zones, the deceased cases in the case of Bhusaval (61 out of 891) are higher than that of Jalgaon MNC (100 out of 2835) as of July end. The main reason behind this scenario can be a higher population density of Bhusaval (18436 /sq.km) than Jalgaon MNC (6788 /sq.km). Lack of social distancing practices, higher illiteracy compared to Jalgaon city, dominant population, delayed reporting of symptoms are some of the reasons behind the higher fatalities in Bhusaval. This zone might have uncontrolled mutations along with moderate medications, which caused higher deaths.

KDMC in the Mumbai metropolitan region is the worst affected containment zone taken under consideration in this work. The total number of COVID-19 positive cases till July 31, 2020, was around 20000, with 357 deceased cases. KDMC has a mixed population scenario in which people in different communities have different occupational practices, diet patterns, housing natures etc. This resulted in higher uncontrolled mutations and recombination of viral strains. Evidence of which may be considered as delay in flattening of curves for daily positive cases reported and active cases. Higher uncontrolled mutations, as well as more spread of infection, might be the reasons behind the higher number of reported deaths.

Through the discussion for deaths and mortality therefore, each containment zone (considered under present work) has its rationale behind the number of deaths that significantly different from the others. To cut a long story short, the overall mortality scenario may be summarized, as shown in Table 6.

**Table 6:** Summary of mortality.

| **Zone** | **Cases / deaths** | **Pop. Density** | **Mortality rationale** |
| --- | --- | --- | --- |
| Mumbai G North | 6750 / 439 | 25358 | • Continuous uncontrolled mutations and recombination.<br>• No physical boundary separation.<br>• More attention to Dharavi, neglecting the rise in cases in other parts. |
| Mumbai M East | 4111 / 292 | 25358 | • Delayed attention from local authorities.<br>• Uncontrolled mutations and recombination.<br>• Lack of counseling.<br>• No physical boundary separation. |
| Kamothe | 1373 / 38 | 14792 | • Uncontrolled mutations.<br>• Mixed population.<br>• No community isolation. |
| KDMC | 19967 / 357 | 18597 | • Uncontrolled mutations.<br>• Mixed population.<br>• Over time the population became weak, resulting in more deaths. |
| Malegaon MNC | 1307 / 85 | 19021 | • Successful community isolation.<br>• Controlled mutations and early saturation of viral strain.<br>• Delayed reporting due to overconfident nature of majority people.<br>• Lack of hospitalization facilities in the initial period. |
| Jalgaon MNC | 2835 / 100 | 6768 | • Mixed population.<br>• Medium hospitalization facilities. |
| Bhusaval | 891 / 61 | 18436 | • Uncontrolled mutations. |
|  |  |  | • Mixed population.<br>• Delayed reporting.<br>• Lack of hospitalization facilities. |
| Sataranjipura | 731 / 8 | 11052 | • Controlled mutations.<br>• Saturation of viral strain in very short period.<br>• Availability of medical facilities from initial period.<br>• Systematic attention and handling by local authorities. |
| Mominpura | 276 / 5 | 11052 |  |
| Solapur MNC | 4985 / 351 | 5345 | • Diet factor dominant in higher deaths.<br>• Lack of high-end hospitalization facilities.<br>• Fear and anxiety. |

To evaluate mortality scenario more effectively, each containment zone (considered under present work) is graphically analyzed for the mortality against the period in the number of days. Mortality, here is considered via different parameter. Cumulative (Cum.) deaths over cumulative (Cum.) period (from starting date considered in study to date of interest in the span of 10 days) are considered as a variable which has direct relation with mortality. This variable is plotted against period. Fits for different functions are obtained and R-square values for all the zones are tabulated, as shown in the following Table 7.

**Table 7:** Fits for mortality.

| Region | Function | R-square |
| --- | --- | --- |
| Kamothe | Exponential | 0.893 |
|  | Logarithmic | 0.833 |
|  | Power | 0.820 |
|  | Linear | 0.894 |
|  | Poly. Second Order | 0.894 |
|  | Poly. Third Order | 0.894 |
|  | Poly. Fourth Order | 0.894 |
|  | Poly. Fifth Order | 1.0 |
|  | Poly. Sixth order | 1.0 |
| KDMC | Exponential | 0.959 |
|  | Logarithmic | 0.906 |
|  | Power | 0.839 |
|  | Linear | 0.956 |
|  | Poly. Second Order | 0.966 |
|  | Poly. Third Order | 0.983 |
|  | Poly. Fourth Order | 0.984 |
|  | Poly. Fifth Order | 0.987 |
|  | Poly. Sixth order | 0.995 |
|  | Exponential | 0.928 |
|  | Logarithmic | 0.851 |
|  | Power | 0.854 |
|  | Linear | 0.873 |
|  | Poly. Second Order | 0.901 |
|  | Poly. Third Order | 0.919 |
|  | Poly. Fourth Order | 0.934 |
|  | Poly. Fifth Order | 0.952 |
|  | Poly. Sixth order | 0.993 |
| Bhusaval | Exponential | 0.497 |
|  | Logarithmic | 0.260 |
|  | Power | 0.281 |
|  | Linear | 0.454 |
|  | Poly. Second Order | 0.615 |
|  | Poly. Third Order | 0.644 |
|  | Poly. Fourth Order | 0.698 |
|  | Poly. Fifth Order | 0.916 |
|  | Poly. Sixth order | 0.999 |
| Sataranjipura | Exponential | 0.231 |
|  | Logarithmic | 0.556 |
|  | Power | 0.420 |
|  | Linear | 0.376 |
|  | Poly. Second Order | 0.799 |
|  | Poly. Third Order | 0.803 |
|  | Poly. Fourth Order | 0.861 |
|  | Poly. Fifth Order | 0.945 |
|  | Poly. Sixth order | 0.995 |
| Mominpura | Exponential | 0.272 |
|  | Logarithmic | 0.145 |
|  | Power | 0.105 |
|  | Linear | 0.328 |
|  | Poly. Second Order | 0.744 |
|  | Poly. Third Order | 0.752 |
|  | Poly. Fourth Order | 0.893 |
|  | Poly. Fifth Order | 0.897 |
|  | Poly. Sixth order | 0.972 |
| Malegaon MNC | Exponential | 0.243 |
|  | Logarithmic | 0.205 |
|  | Power | 0.230 |
|  | Linear | 0.212 |
|  | Poly. Second Order | 0.220 |
|  | Poly. Third Order | 0.300 |
|  | Poly. Fourth Order | 0.316 |
|  | Poly. Fifth Order | 0.658 |
|  | Poly. Sixth order | 0.960 |
| Solapur MNC | Exponential | 0.679 |
|  | Logarithmic | 0.432 |
|  | Power | 0.471 |
|  | Linear | 0.618 |
|  | Poly. Second Order | 0.655 |
|  | Poly. Third Order | 0.749 |
|  | Poly. Fourth Order | 0.940 |
|  | Poly. Fifth Order | 0.998 |
|  | Poly. Sixth order | 0.998 |

As evident from the above Table, R-square values for mortality against the number of days with polynomial functions are statistically significant. Hence for brevity, graphs of the mortality for polynomial functions are plotted as shown in Fig.31 through Fig.38. Polynomial of sixth order mimic the behaviour of cumulative deaths per cumulative infections. (From the beginning date till the date of interest). It can be noted that for most of the containment zones, deaths are reducing though by wavy manner, in particular for polynomials of order 6. Further corresponding fits are reasonable as well. For Mominpura, though trend is slightly different as in Fig.33, still as deaths reported are less, this is not a cause of concern. Kamothe zone is not slowing down quickly and aftershocks will continue possibly due to multiple reasons outlined in preceding sections.

**Fig.31.**
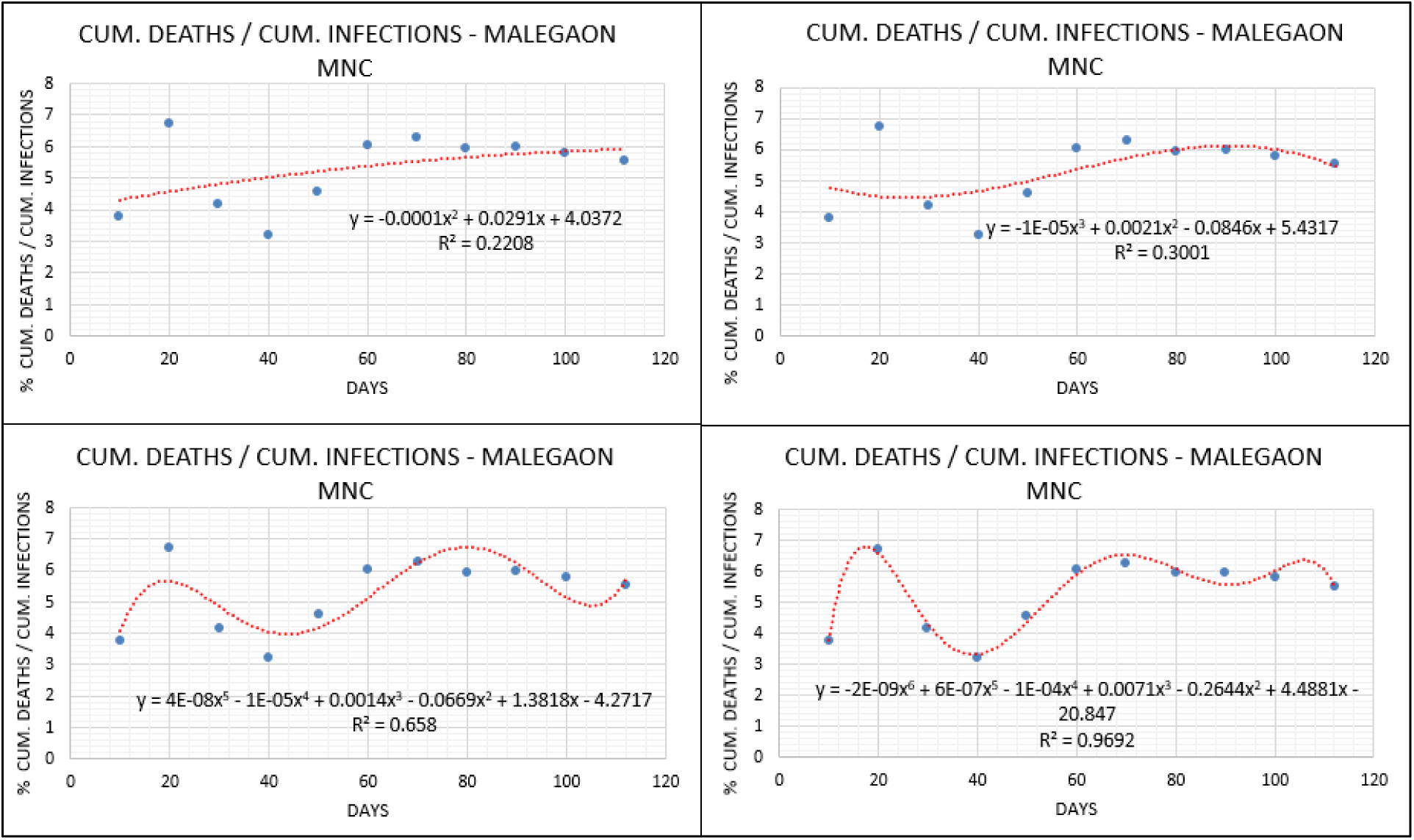
Cum. deaths / cum. infections vs. Period – Malegaon MNC. (Cum. deaths and Cum. infections are taken from the starting date to the date of interest over the period of 10 days by cum. manner)

**Fig.32.**
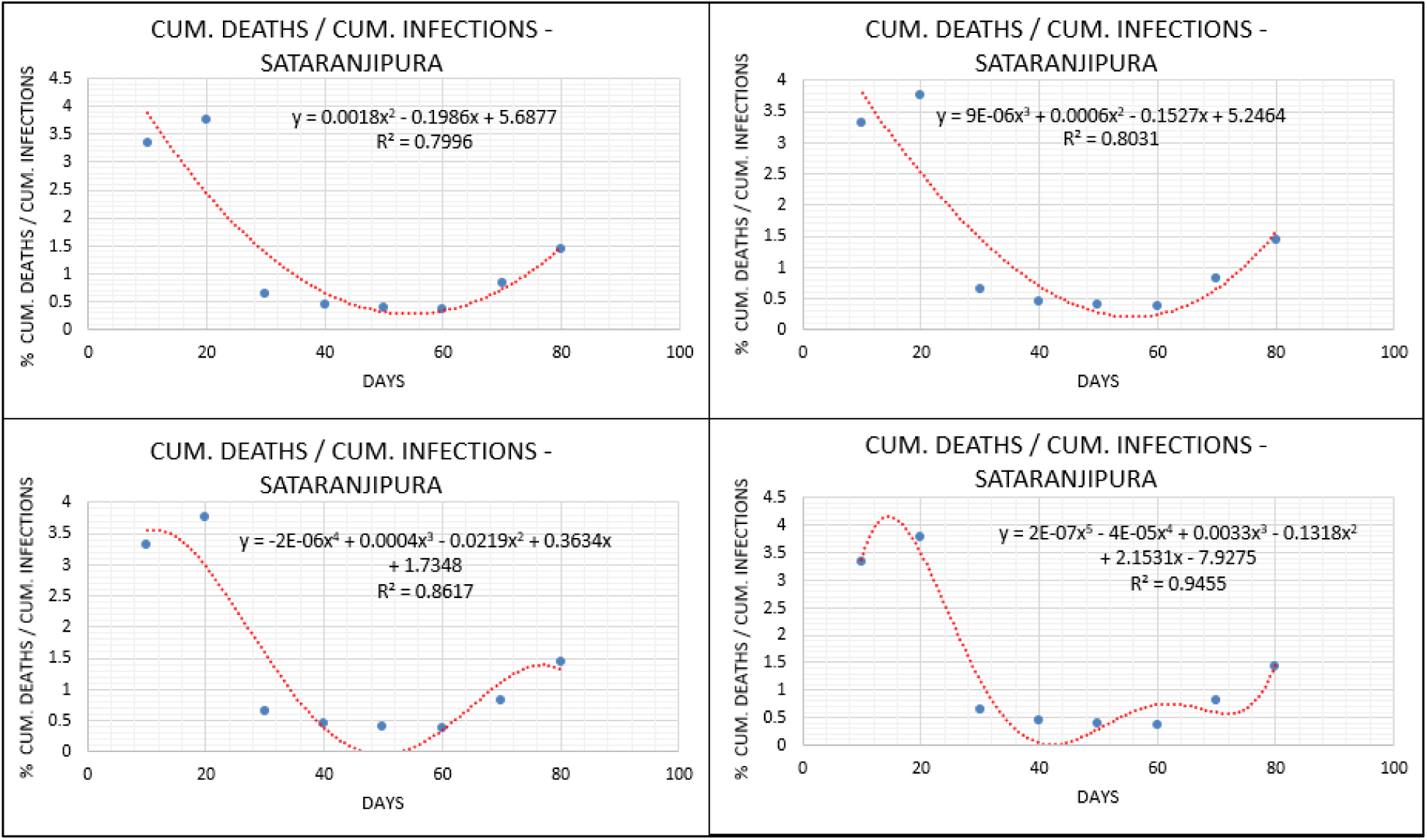
Cum. deaths / Cum. infections vs. Period – Sataranjipura. (Cum. deaths and Cum. infections are taken from the starting date to the date of interest over the period of 10 days by cum. manner)

**Fig.33.**
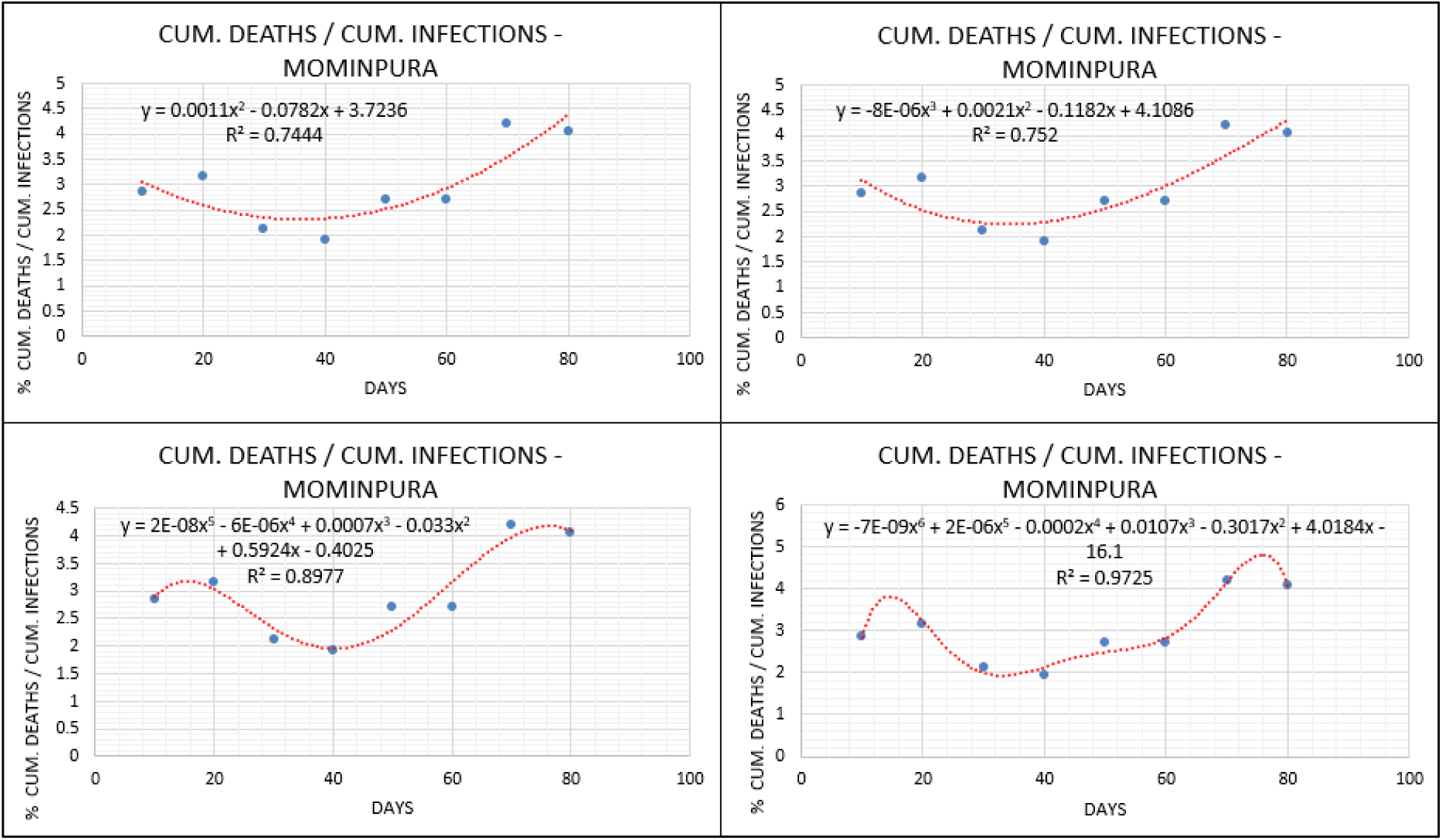
Cum. deaths / Cum. infections vs. Period – Mominpura. (Cum. deaths and Cum. infections are taken from the starting date to the date of interest over the period of 10 days by cum. manner)

**Fig.34.**
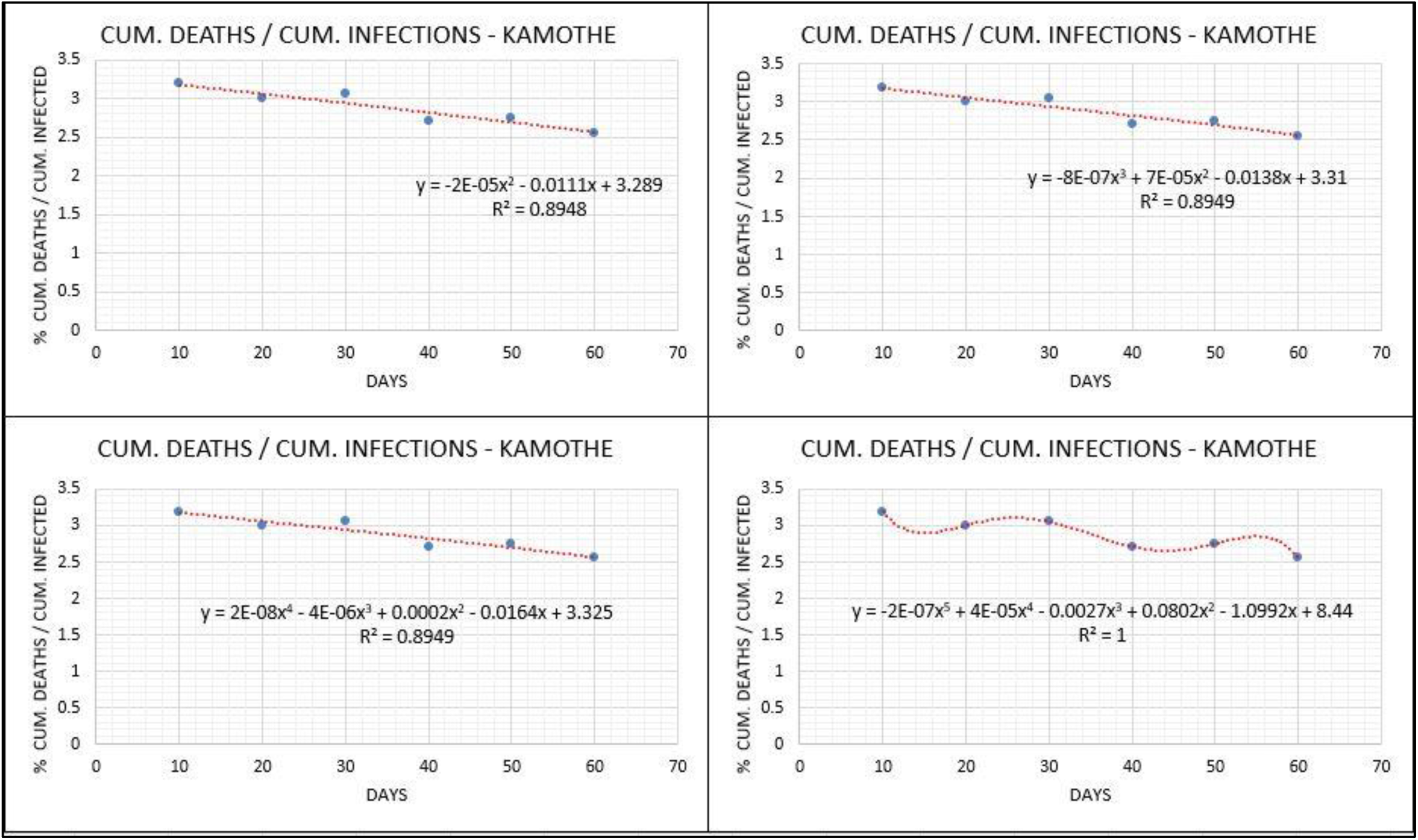
Cum. deaths / Cum. infections vs. Period – Kamothe. (Cum. deaths and Cum. infections are taken from the starting date to the date of interest over the period of 10 days by cum. manner)

**Fig.35.**
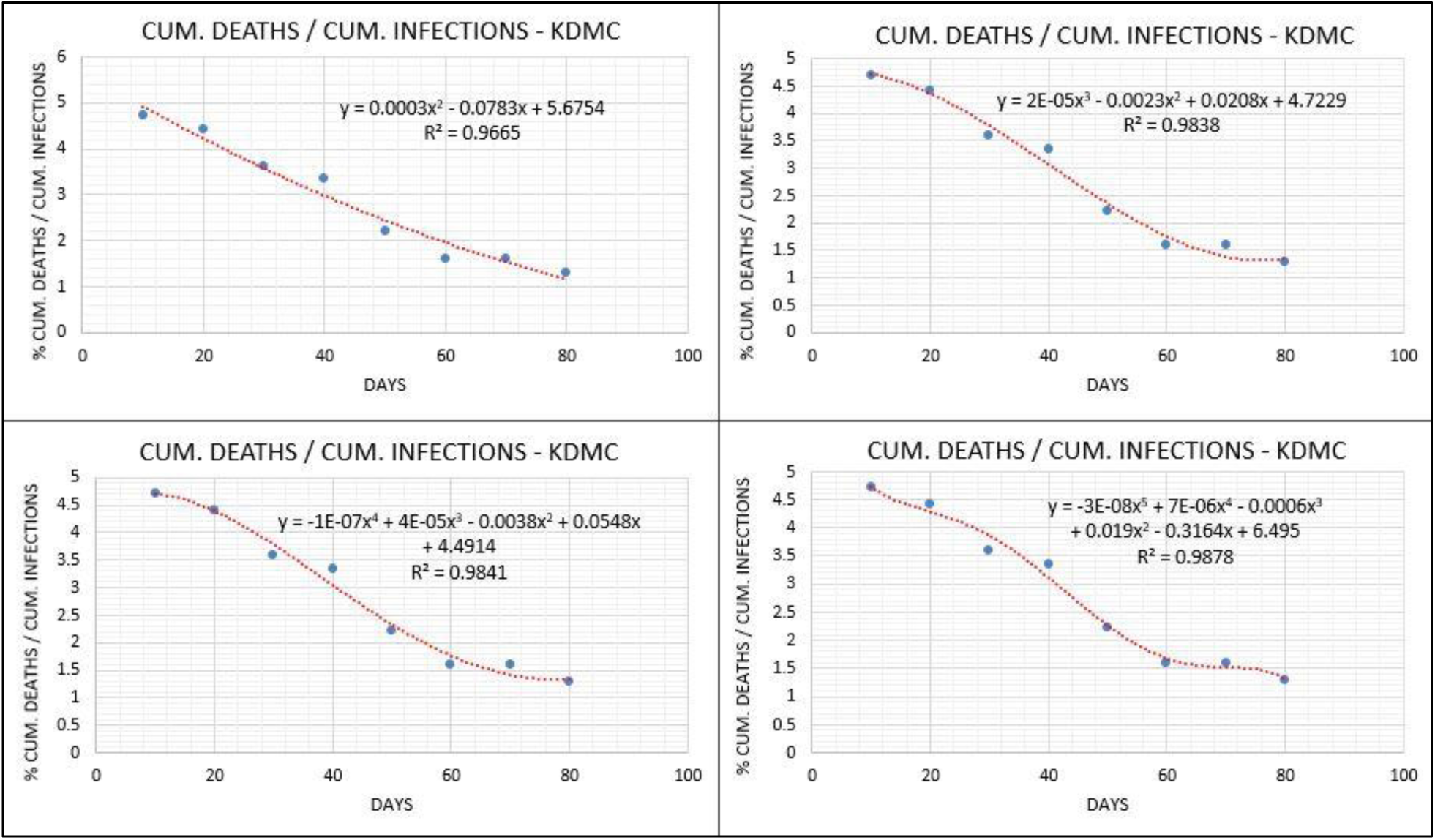
Cum. deaths / Cum. infections vs. Period – KDMC. (Cum. deaths and Cum. infections are taken from the starting date to the date of interest over the period of 10 days by cum. manner)

**Fig.36.**
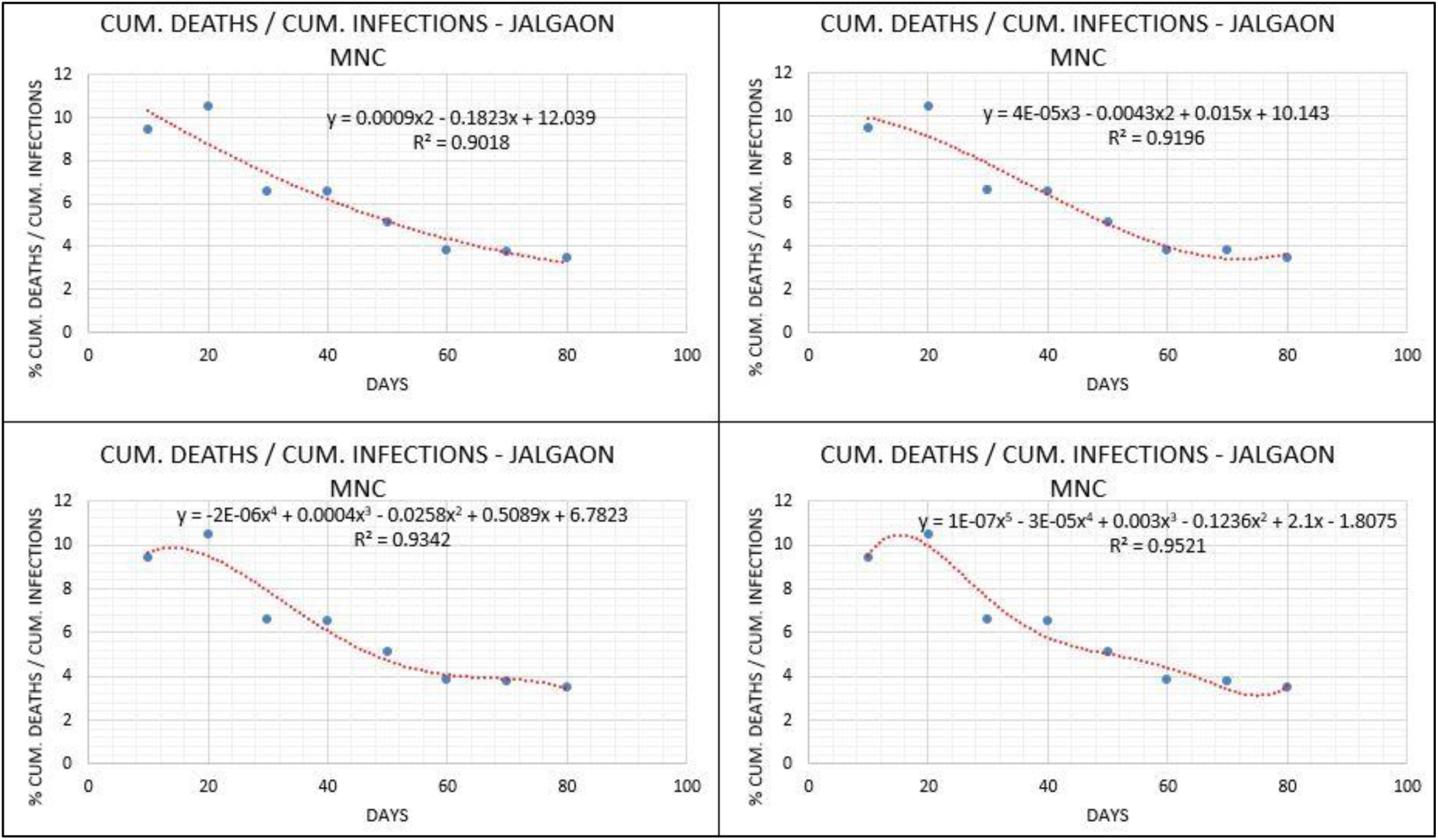
Cum. deaths / Cum. infections – Jalgaon MNC. (Cum. deaths and Cum. infections are taken from the starting date to the date of interest over the period of 10 days by cum. manner)

**Fig.37.**
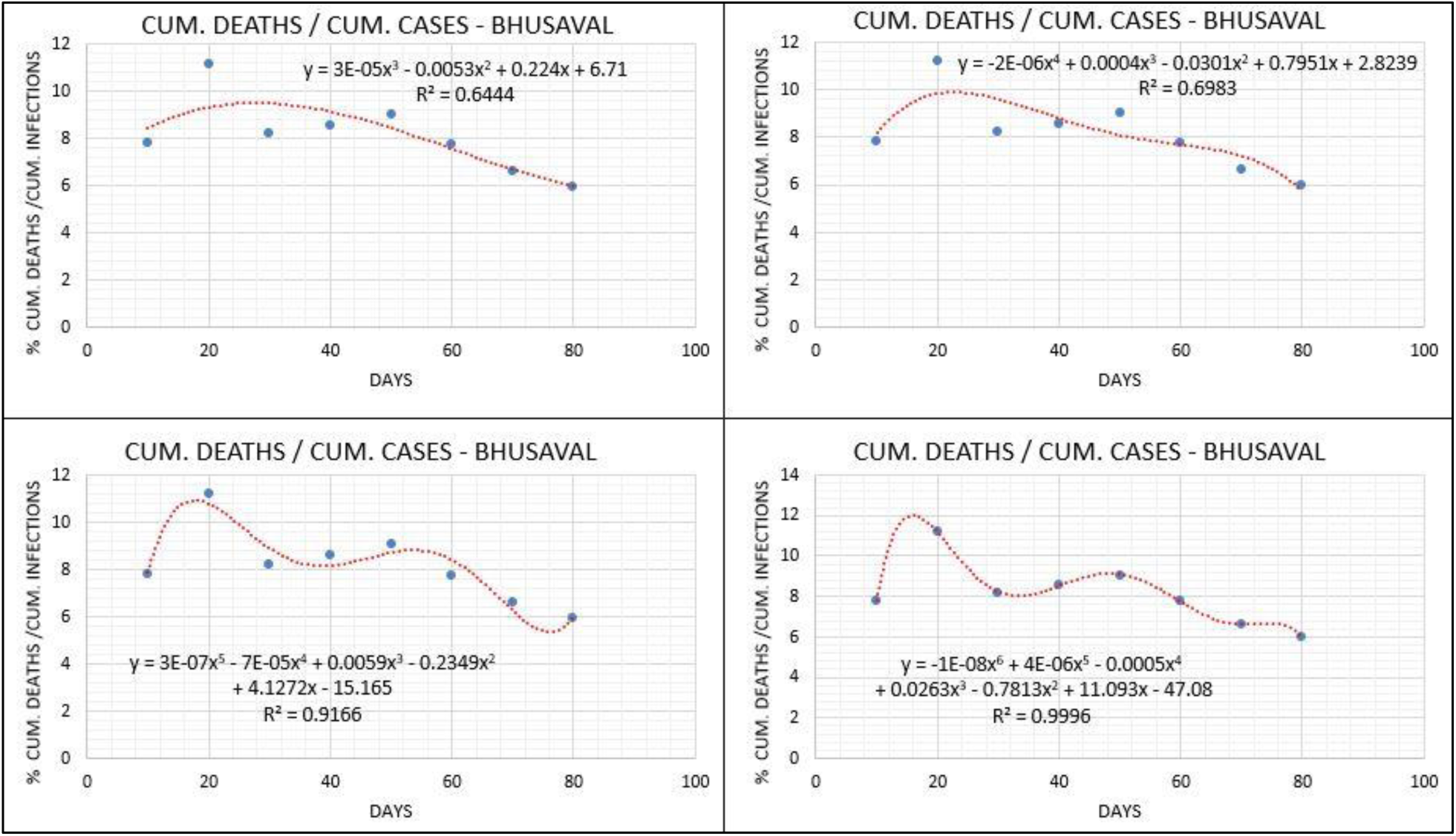
Cum. deaths / Cum. infections – Bhusaval. (Cum. deaths and Cum. infections are taken from the starting date to the date of interest over the period of 10 days by cum. manner)

**Fig.38.**
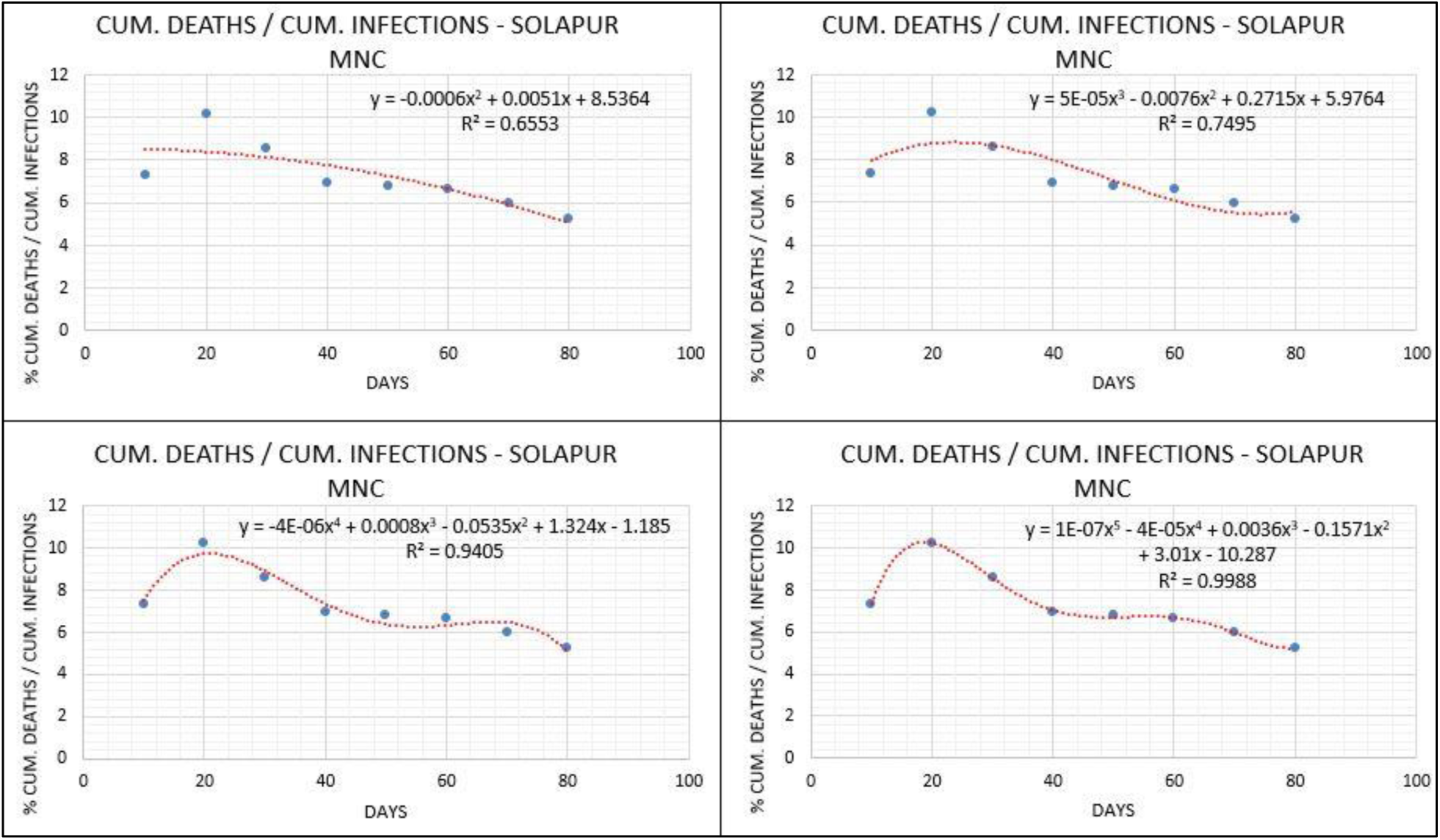
Cum. deaths / Cum. infections – Solapur MNC. (Cum. deaths and Cum. infections are taken from the starting date to the date of interest over the period of 10 days by cum. manner)

## 4. Conclusion

Critical containment zones in Maharashtra are analyzed in the context of daily infected cases, cumulative infected cases, recovered cases, cumulative recovered cases, active (net) cases and deaths. Below are the key findings of the work.

- Polynomial of order six mimic the wavy nature of COVID-19 critical as well as general outcomes. Pandemic will not vanish in Maharashtra and similar demographies at least for some unpredictable time.
- Statistical analysis shown lower R-square for daily reported cases, whereas significant insights are produced when cumulative cases are considered.
- Hence it is advisable to consider the cumulative data in computations of the viral outbreaks like the COVID-19.
- Large scale recoveries and a significant reduction in the active cases are observed for mean temperature range of 26℃ to 30℃ and mean RH range of 70% to 85%.
- In non-coastal regions, different transmission mechanism exists because of different aerosol size which is responsible for symptomatic cases. On the other hand coastal regions do show larger aerosol responsible for more asymptomatic cases.
- Behavioural factors such as fear, anxiety and stress are controlling COVID-19 outcomes.
- Areas having a dominant population showed controlled mutations and early saturation. On the other hand, areas having a mixed population faced uncontrolled mutations as well as recombination.
- Intake of essential food content changes according to social demography. Abrupt change in dietary patterns resulted in comorbidities.
- Each containment zone considered, has a rationale for mortality as well as infection patterns, which is unique than the others. This is important for scientists, researchers, pharmacists and biotechnologists in the context of developing the vaccine for COVID-19 and administrating them over the people from different localities of Maharashtra with more emphasized testing on both larger size of cohorts along with large numbers of such cohorts.
- Government authorities can plan for the deployment of specialized healthcare units for controlling localized outbreaks as lack of high-end medical facilities in rural areas which resulted in high mortality.

We highlight the need for the moderate exercise with balanced diet, reduced fear, stress and anxiety concerns, reduction in natural disturbances, better medications at rural locations etc. to control COVID-19 outcomes and post COVID-19 era. Further we encourage other researchers to emphasize the study involving the transmission patterns among various blood groups using matrix method and abreast AI framework. In a nutshell, as the groundwork part, the policymakers and vaccine developers must consider the areas like the state of Maharashtra across globe, which have high divergence in the context of COVID-19 problems and remedies. This will help in acquiring precise solutions quickly without the adverse effects (Post COVID-19).

## Data Availability

https://twitter.com/InfoNashik,
https://twitter.com/InfoJalgaon,
https://twitter.com/ngpnmc,
https://twitter.com/PanvelCorp,
https://twitter.com/Maha_MEDD,
https://www.ncbi.nlm.nih.gov/books/NBK8439/

## Author Contributions

All author contributions are equal.

